# QCovid 4 - Predicting risk of death or hospitalisation from COVID-19 in adults testing positive for SARS-CoV-2 infection during the Omicron wave in England

**DOI:** 10.1101/2022.08.13.22278733

**Authors:** Julia Hippisley-Cox, Kamlesh Khunti, Aziz Sheikh, Jonathan S Nguyen-Van-Tam, Carol AC Coupland

## Abstract

**Objectives:** To (a) derive and validate risk prediction algorithms (QCovid4) to estimate risk of COVID-19 mortality and hospitalisation in UK adults with a SARS-CoV-2 positive test during the ‘Omicron’ pandemic wave in England and (b) evaluate performance with earlier versions of algorithms developed in previous pandemic waves and the high-risk cohort identified by NHS Digital in England.

**Design:** Population-based cohort study using the QResearch database linked to national data on COVID-19 vaccination, high risk patients prioritised for COVID-19 therapeutics, SARS-CoV-2 results, hospitalisation, cancer registry, systemic anticancer treatment, radiotherapy and the national death registry.

**Settings and study period:** 1.3 million adults in the derivation cohort and 0.15 million adults in the validation cohort aged 18-100 years with a SARS-CoV-2 positive test between 11^th^ December 2021 and 31^st^ March 2022 with follow up to 30^th^ June 2022.

**Main outcome measures:** Our primary outcome was COVID-19 death. The secondary outcome of interest was COVID-19 hospital admission. Models fitted in the derivation cohort to derive risk equations using a range of predictor variables. Performance evaluated in a separate validation cohort.

**Results:** Of 1,297,984 people with a SARS-CoV-2 positive test in the derivation cohort, 18,756 (1.45%) had a COVID-19 related hospital admission and 3,878 (0.3%) had a COVID-19 death during follow-up. Of the 145,404 people in the validation cohort, there were 2,124 (1.46%) COVID-19 admissions and 461 (0.3%) COVID-19 deaths.

The COVID-19 mortality rate in men increased with age and deprivation. In the QCovid4 model in men hazard ratios were highest for those with the following conditions (for 95% CI see Figure 1): kidney transplant (6.1-fold increase); Down’s syndrome (4.9-fold); radiotherapy (3.1-fold); type 1 diabetes (3.4-fold); chemotherapy grade A (3.8-fold), grade B (5.8-fold); grade C (10.9-fold); solid organ transplant ever (2.4-fold); dementia (1.62-fold); Parkinson’s disease (2.2-fold); liver cirrhosis (2.5-fold). Other conditions associated with increased COVID-19 mortality included learning disability, chronic kidney disease (stages 4 and 5), blood cancer, respiratory cancer, immunosuppressants, oral steroids, COPD, coronary heart disease, stroke, atrial fibrillation, heart failure, thromboembolism, rheumatoid/SLE, schizophrenia/bipolar disease sickle cell/HIV/SCID; type 2 diabetes. Results were similar in the model in women.

COVID-19 mortality risk was lower among those who had received COVID-19 vaccination compared with unvaccinated individuals with evidence of a dose response relationship. The reduced mortality rates associated with prior SARS-CoV-2 infection were similar in men (adjusted hazard ratio (HR) 0.51 (95% CI 0.40, 0.64)) and women (adjusted HR 0.55 (95%CI 0.45, 0.67)).

The QCOVID4 algorithm explained 76.6% (95%CI 74.4 to 78.8) of the variation in time to COVID-19 death (R^2^) in women. The D statistic was 3.70 (95%CI 3.48 to 3.93) and the Harrell’s C statistic was 0.965 (95%CI 0.951 to 0.978). The corresponding results for COVID-19 death in men were similar with R^2^ 76.0% (95% 73.9 to 78.2); D statistic 3.65 (95%CI 3.43 to 3.86) and C statistic of 0.970 (95%CI 0.962 to 0.979). QCOVID4 discrimination for mortality was slightly higher than that for QCOVID1 and QCOVID2, but calibration was much improved.

**Conclusion:** The QCovid4 risk algorithm modelled from data during the UK’s Omicron wave now includes vaccination dose and prior SARS-CoV-2 infection and predicts COVID-19 mortality among people with a positive test. It has excellent performance and could be used for targeting COVID-19 vaccination and therapeutics. Although large disparities in risks of severe COVID-19 outcomes among ethnic minority groups were observed during the early waves of the pandemic, these are much reduced now with no increased risk of mortality by ethnic group.

**What is known:**

- The QCOVID risk assessment algorithm for predicting risk of COVID-19 death or hospital admission based on individual characteristics has been used in England to identify people at high risk of severe COVID-19 outcomes, adding an additional 1.5 million people to the national shielded patient list in England and in the UK for prioritising people for COVID-19 vaccination.
- There are ethnic disparities in severe COVID-19 outcomes which were most marked in the first pandemic wave in 2020.
- COVID-19 vaccinations and therapeutics (monoclonal antibodies and antivirals) are available but need to be targeted to those at highest risk of severe outcomes.

**What this study adds:**

- The QCOVID4 risk algorithm using data from the Omicron wave now includes number of vaccination doses and prior SARS-CoV-2 infection. It has excellent performance both for ranking individuals (discrimination) and predicting levels of absolute risk (calibration) and can be used for targeting COVID-19 vaccination and therapeutics as well as individualised risk assessment.
- QCOVID4 more accurately identifies individuals at highest levels of absolute risk for targeted interventions than the ‘conditions-based’ approach adopted by NHS Digital based on relative risk of a list of medical conditions.
- Although large disparities in risks of severe COVID-19 outcomes among ethnic minority groups were observed during the early waves of the pandemic, these are much reduced now with no increased risk of mortality by ethnic group.

## INTRODUCTION

During the first waves of the COVID-19 pandemic, before the introduction of vaccines, it was essential to be able to identify people at highest risk of severe COVID-19 outcomes if they were infected with SARS-CoV-2 virus. The QCOVID risk assessment tool for predicting risk of COVID-19 death or hospital admission based on individual characteristics was developed,^1^ independently externally validated in England,^2^ Wales^3^ and Scotland,^4^ and found to have excellent performance for identifying those at high risk of severe outcomes from COVID-19. QCOVID was used in England in February 2021 to identify patients at high risk of severe COVID-19 outcomes, adding an additional 1.5 million people to the national shielded patient list. It was also used for prioritising people for vaccination across the UK (if they had not already been offered the vaccine on account of their age or other risk classification)^5^. The QCOVID model was updated following the second and third waves of the pandemic to create two new versions of the model, QCOVID2 based on unvaccinated patients^6^ and QCOVID3 based on partially vaccinated patients.^6^ These models accounted for changes that had occurred both in the virus as well as the deployment of the vaccination programme.^6^

In December 2021, the UK experienced a new wave of COVID-19 infections with the Omicron variant rapidly replacing high circulating levels of the previous Delta variant. Whilst the Omicron variant (BA1) was associated with a lower risk of COVID-19 death than the Delta variant,^7^ further mutations have occurred and there are concerns that COVID-19 vaccines may become less effective. Additional therapeutic agents are likely to be needed to protect vulnerable individuals such as antivirals and neutralising monoclonal antibodies (nMABS^8^). On 9^th^ December 2021, nMABs became available in the UK for high-risk non-hospitalised patients with a symptomatic SARS-CoV-2 infection.^9^ Since nMABs are limited resources, they have been targeted to those at highest risk of poor COVID-19 outcomes who are most likely to benefit.^9 10^ This was done based on a set of clinical conditions associated with a high relative risk of severe outcomes from the published literature^6 11^ combined with clinical judgement regarding likelihood of clinical benefit based on the biological mechanism for nMABS ^9^. Patients with these conditions were then identified from centrally held electronic health records and contacted by NHS Digital in December 2021 to inform them of their potential eligibility for nMABS should they develop a symptomatic SARS-CoV-2 infection. The guidance did not however, account for the cumulative absolute risk associated with multiple co-morbidities, age, prior infection, vaccination status or the new variants.

The aims of this study, commissioned by the UK’s Department of Health and Social Care, were to develop and validate a new QCOVID risk algorithm (QCOVID 4) based on new data from the Omicron pandemic wave in England, accounting for prior SARS-CoV-2 infection and number of COVID-19 vaccination doses. We also evaluated the performance of the QCOVID4 algorithm with earlier versions of the risk model developed in the first two pandemic waves and the ‘high risk’ cohort identified by NHS Digital on the basis of relative risk of a list of conditions. The results can then be used to inform ongoing strategies for targeting therapeutics and other public health interventions, designed to protect those most at risk from COVID-19 death and hospitalisation.

## METHODS

### Data sources

We used the QResearch database (version 47) of 12 million current patients with demographic, clinical and medication data which is used for epidemiological^12 1^ and drug safety research.^13 14^ QResearch is linked to multiple datasets at individual patient level. For this analysis, we used the following linked datasets

- National Immunisation (NIMS) Database of COVID-19 vaccinations to identify data on vaccine dates and doses for all people vaccinated in England
- Hospital Episode Statistics (HES) dataset supplemented by the more regularly updated Secondary Users Service data (SUS-PLUS).
- Civil registration national data for mortality with date and up to 15 causes of death
- SARS-CoV-2 infection data, Second Generation Surveillance System (SGSS) and Pillar 2)
- Systemic anticancer treatment (SACT) data
- NHS Digital ‘high risk’ cohort prioritised for novel COVID-19 therapeutics in December 2021.

### Study design and period for cohort

We undertook a cohort study of all individuals aged 18-100 years who had one or more positive SARS-CoV-2 tests from 11 December 2021 (the date of the first notified Omicron case) to 31 March 2022 (the date after which widespread free NHS SARS-CoV-2 tests became unavailable). Individuals were followed from the date of their first SARS-CoV-2 test in the study period, until they had the outcome of interest, died or the end of the study period on 30 June 2022 (the latest date for which mortality and hospital admissions were available).

### Outcomes for cohort

The primary outcome was time to COVID-19 death (either in hospital or out of hospital) as recorded in any position on the death certificate or death within 28 days of a SARS-CoV-2 positive test. The secondary outcome was time to hospital admission with COVID-19, defined as either confirmed or suspected COVID-19 on International Classification of Diseases (ICD)-10 code (U071, U072). We used these definitions for the outcomes for consistency with other QCovid algorithms and because these are the ones used for COVID-19 death and hospital admission in the UK.^15^

### Predictor variables

Candidate predictor variables were those previously identified as associated with increased risk of COVID-19 death or hospitalisation from the original QCOVID protocol^16^ and the published literature^1 6 12 17^. The variables were: age, sex, ethnicity, Townsend material deprivation (an area level score based on postcode where higher scores indicate higher levels of deprivation ^18^), number of vaccine doses (none, 1, 2, 3, 4 or more), body mass index (BMI) ^17^, domicile (care home, homeless, neither); chronic kidney disease (CKD); chemotherapy in previous 12 months; type 1 or type 2 diabetes (with glycosylated haemoglobin, HbA1C <59 or ≥59 mmol/mol); blood cancer; bone marrow transplant in last six months; respiratory cancer; radiotherapy in last six months; solid organ transplant; chronic obstructive pulmonary disease (COPD); asthma; rare lung diseases (cystic fibrosis, bronchiectasis or alveolitis); pulmonary hypertension or pulmonary fibrosis); coronary heart disease; stroke; atrial fibrillation; heart failure; venous thromboembolism; peripheral vascular disease; congenital heart disease; dementia; Parkinson’s disease; epilepsy; Down’s syndrome; rare neurological conditions (motor neurone disease, multiple sclerosis, myasthenia or Huntington’s Chorea); cerebral palsy; osteoporotic fracture; rheumatoid arthritis or systemic lupus erythematosus (SLE); liver cirrhosis; bipolar disorder or schizophrenia; inflammatory bowel disease; sickle cell disease; Human Immunodeficiency Virus (HIV) or Acquired Immunodeficiency Syndrome (AIDS); and Severe Combined Immunodeficiency (SCID).

We defined predictors using information recorded in primary care electronic health records at the start of follow-up (date of first positive SARS-CoV-2 test in the study period), except for data for the number of SARS-CoV-2 infections, COVID-19 vaccinations, chemotherapy, radiotherapy and transplants, which were based on linked secondary care data. For all predictor variables, we used the most recently available value at the cohort entry date.

### Model development

As in previous studies, we used 90% of practices to develop the models and the remaining 10% of practices for model validation.^6^ We developed separate risk models in men and women using Cox proportional hazard models to calculate hazard ratios (HRs) for the two outcomes. We used second degree fractional polynomials to model non-linear relationships for continuous variables including age, BMI and Townsend material deprivation score.^18^ We used multiple imputation with chained equations to impute missing values for ethnicity, Townsend score, BMI and HBA1C. We carried out five imputations and fitted the prediction models in each imputed dataset. We used Rubin’s rules^19^ to combine the model parameter estimates across the imputed datasets.

We retained variables in the final models that were significant at the 5% level and where adjusted hazard ratios were > 1.1. We combined clinically similar variables with very low numbers of events. We examined interactions between predictor variables and age. We estimated the baseline survivor function based on zero values of centred continuous variables, with all binary predictor values set to zero. We used the regression coefficients for each variable from the final model as weights which we combined with the baseline survivor function evaluated at 30, 60 and 90 days of follow-up.^20^

### Model evaluation

We evaluated model performance in the validation cohort. We used multiple imputation to replace missing values for ethnicity, HBA1C, BMI and Townsend score using the same imputation model as in the derivation cohort. We applied the final risk equations to calculate the risk scores for each outcome. We calculated Harrell’s C statistics ^21^, R^2^ values and D statistics^22^. We assessed model calibration in the validation cohort by comparing mean predicted risks at 90 days with the observed risks by twentieths of predicted risk^23^.

We calculated each performance metric in the whole validation cohort and in subgroups for age and ethnic group (where numbers allowed). We compared model discrimination with risk scores calculated using earlier versions of QCOVID developed on unvaccinated populations:

a. during the first pandemic wave (QCOVID1, developed on the total unvaccinated population between 24 January 2020 and 30 April 2020) and
b. during the second pandemic wave (QCOVID2, developed on the unvaccinated patients with a SARS-CoV-2 positive test between 8 December 2020 to 21 June 2021 during which the Alpha and Delta variants were dominant).

We decided not to include QCOVID3 as that was developed on a partially vaccinated population during the rapid roll-out of the vaccination program. We also determined whether there were identifiable high-risk groups based on their QCOVID4 predicted risks who had an equivalent or higher observed risk of COVID-19 death than the current ‘high risk cohort’ by comparing groups of the same size (high risk vs QCOVID4 threshold).

### Risk stratification

We applied the QCOVID4 algorithms to the validation cohort to define the centile thresholds based on absolute predicted risk. We calculated sensitivity as the total number of patients with a risk score above the risk threshold with a COVID-19 death out of the total number of COVID-19 deaths.

We also compared risk stratification using (a) QCOVID4 to identify the top 2.5% of patients at highest absolute risk in the validation cohort with (b) the current recommended guidelines which have selected a high-risk cohort based on relative risk of patients with selected medical conditions.

### Reporting

We adhered to the RECORD^24^ and TRIPOD statements for reporting ^25^. We used all the available data on the database to maximise the power and generalisability of the results. We used STATA (version 17) for analyses.

#### Patient and public involvement

Patients were involved in framing research question, identifying predictors and in developing plans for design and implementation of the QCOVID risk tool. A citizen’s jury convened by the Scottish government, evaluated earlier versions of the QCovid algorithm and highlighted the importance of keeping it up-to-date and maintaining transparency over its use ^26^. Patients will be invited to advise on disseminating the results including the development of infographics and its translation into different languages.

## RESULTS

### Baseline characteristics of study cohorts

Overall, there were 1,430 practices in the QResearch database (version 47). We allocated 1,287 practices to the derivation cohort and 143 to the validation cohort. Of the 9,526,580 patients aged 18-100 years in the derivation cohort, 1,297,922 (13.6%) had a SARS-CoV-2 positive test in the study period. Of these, 18,756 (1.5%) had a COVID-19 hospital admission and 3878 (0.3%) had a COVID-19 death during follow-up.

Of the 1,064,255 patients in the validation cohort, 145,397 (13.7%) had a SARS-CoV-2 positive test and were included in the analysis. Of these, 2,124 (1.5%) had a COVID-19 hospital admission and 461 (0.3%) had a COVID-19 death.

Table 1 shows the baseline characteristics of those who tested positive, those with a COVID-19 death and those with a COVID-19 admission in the derivation cohort. Supplementary table 1 shows the corresponding results for the validation cohort. The mean age in the derivation cohort for those with a SARS-CoV-2 positive test was 42.4 years (SD 16.4), COVID-19 admission 55.6 years (SD 22.1), COVID-19 death, 80.9 years (SD 12.3)

**TABLE 1.** Baseline characteristics of the derivation cohort of patients with a SARS-CoV-2 positive test, COVID-19 death and COVID-19 admission. (*cells with counts <5 suppressed)

|  | *total population* | *SARS-CoV-2*<br>*positive* | *COVID-19 death* | *COVID-19*<br>*admission* |
| --- | --- | --- | --- | --- |
| total | 9,526,580 | 1,297,922 | 3,878 | 18,756 |
| males (%) | 4776225 (50.14) | 555918 (42.83) | 2026 (52.24) | 7708 (41.10) |
| mean age (SD) | 47.22 (18.57) | 42.39 (16.44) | 80.93 (12.29) | 55.63 (22.13) |
| White | 6071398 (63.73) | 883218 (68.05) | 2944 (75.92) | 12913 (68.85) |
| Indian | 302042 (3.17) | 35493 (2.73) | 49 (1.26) | 411 (2.19) |
| Pakistani | 183637 (1.93) | 16780 (1.29) | 40 (1.03) | 405 (2.16) |
| Bangladeshi | 120383 (1.26) | 11411 (0.88) | 28 (0.72) | 253 (1.35) |
| Other Asian | 194823 (2.05) | 23926 (1.84) | 34 (0.88) | 336 (1.79) |
| Caribbean | 104802 (1.10) | 12256 (0.94) | 64 (1.65) | 378 (2.02) |
| Black African | 250975 (2.63) | 25970 (2.00) | 36 (0.93) | 531 (2.83) |
| Chinese | 110390 (1.16) | 8824 (0.68) | 9 (0.23) | 70 (0.37) |
| Other ethnic group | 403014 (4.23) | 51419 (3.96) | 47 (1.21) | 759 (4.05) |
| Ethnicity not recorded | 1785116 (18.74) | 228625 (17.61) | 627 (16.17) | 2700 (14.40) |
| Deprivation quintile |  |  |  |  |
| 1 (most affluent) | 2241175 (23.53) | 313128 (24.13) | 987 (25.45) | 3827 (20.40) |
| 2 | 2030972 (21.32) | 289819 (22.33) | 888 (22.90) | 3764 (20.07) |
| 3 | 1850166 (19.42) | 260461 (20.07) | 785 (20.24) | 3863 (20.60) |
| 4 | 1682062 (17.66) | 220973 (17.03) | 689 (17.77) | 3715 (19.81) |
| 5 (most deprived) | 1603423 (16.83) | 192601 (14.84) | 506 (13.05) | 3358 (17.90) |
| 6 (not recorded) | 118782 (1.25) | 20940 (1.61) | 23 (0.59) | 229 (1.22) |
| neither | 9436601 (99.06) | 1279698 (98.60) | 2997 (77.28) | 17803 (94.92) |
| Care home | 70424 (0.74) | 16703 (1.29) | 876 (22.59) | 873 (4.65) |
| Homeless | 19555 (0.21) | 1521 (0.12) | 5 (0.13) | 80 (0.43) |
| BMI < 18.5 | 239052 (2.51) | 32788 (2.53) | 287 (7.40) | 570 (3.04) |
| BMI 18.5-24.99 | 2770605 (29.08) | 398814 (30.73) | 1251 (32.26) | 5083 (27.10) |
| BMI 25-29.99 | 2339194 (24.55) | 311541 (24.00) | 942 (24.29) | 4736 (25.25) |
| BMI 30-34.99 | 1096337 (11.51) | 146966 (11.32) | 470 (12.12) | 2682 (14.30) |
| BMI 35+ | 425986 (4.47) | 60829 (4.69) | 156 (4.02) | 1223 (6.52) |
| BMI 40+ | 224232 (2.35) | 33922 (2.61) | 82 (2.11) | 729 (3.89) |
| BMI not recorded | 2431174 (25.52) | 313062 (24.12) | 690 (17.79) | 3733 (19.90) |
| no CKD | 9166042 (96.22) | 1267768 (97.68) | 2557 (65.94) | 16082 (85.74) |
| CKD5 only | 10386 (0.11) | 1384 (0.11) | 67 (1.73) | 238 (1.27) |
| CKD5 with dialysis | 3178 (0.03) | 484 (0.04) | 10 (0.26) | 49 (0.26) |
| CKD5 with transplant | 5391 (0.06) | 1123 (0.09) | 26 (0.67) | 251 (1.34) |
| no learning disability | 9353294 (98.18) | 1275513 (98.27) | 3773 (97.29) | 18189 (96.98) |
| Learning disability | 169009 (1.77) | 21745 (1.68) | 102 (2.63) | 529 (2.82) |
| Downs syndrome | 4277 (0.04) | 664 (0.05) | * | 38 (0.20) |
| No chemo in last 12 | 9490847 (99.62) | 1293639 (99.67) | 3633 (93.68) | 18205 (97.06) |
| Chemo group A | 16515 (0.17) | 1683 (0.13) | 76 (1.96) | 169 (0.90) |
| chemo group B | 18150 (0.19) | 2445 (0.19) | 155 (4.00) | 356 (1.90) |
| chemo group C | 1068 (0.01) | 155 (0.01) | 14 (0.36) | 26 (0.14) |
| no type1 diabetes | 9480133 (99.51) | 1290601 (99.44) | 3855 (99.41) | 18491 (98.59) |
| type 1 HBA<=59 | 13894 (0.15) | 2354 (0.18) | 5 (0.13) | 83 (0.44) |
| type1 HBA1C 59+ | 27110 (0.28) | 4136 (0.32) | 14 (0.36) | 149 (0.79) |
| type 1 HBA1C not rec | 5443 (0.06) | 831 (0.06) | * | 33 (0.18) |
| no type2 diabetes | 8901793 (93.44) | 1241431 (95.65) | 2814 (72.56) | 15849 (84.50) |
| type 2 HBA<=59 | 349984 (3.67) | 31490 (2.43) | 639 (16.48) | 1578 (8.41) |
| type2 HBA1C 59+ | 202963 (2.13) | 18246 (1.41) | 288 (7.43) | 929 (4.95) |
| type 2 HBA1C not rec | 71840 (0.75) | 6755 (0.52) | 137 (3.53) | 400 (2.13) |
| no covid vaccine doses | 1719164 (18.05) | 145887 (11.24) | 413 (10.65) | 3720 (19.83) |
| 1 dose | 279909 (2.94) | 44125 (3.40) | 110 (2.84) | 803 (4.28) |
| 2 doses | 1519472 (15.95) | 363949 (28.04) | 670 (17.28) | 3832 (20.43) |
| 3 doses | 5194720 (54.53) | 737212 (56.80) | 2617 (67.48) | 9861 (52.58) |
| 4 doses | 813315 (8.54) | 6749 (0.52) | 68 (1.75) | 540 (2.88) |
| SARS-CoV-2 infection<br>prior to study entry | 1322848 (13.89) | 145587 (11.22) | 177 (4.56) | 1384 (7.38) |
| Blood cancer | 72167 (0.76) | 8389 (0.65) | 261 (6.73) | 741 (3.95) |
| Bone marrow transplant<br>in last 6/12 | 332 (0.00) | 44 (0.00) | * | 11 (0.06) |
| Respiratory cancer | 20163 (0.21) | 1743 (0.13) | 101 (2.60) | 167 (0.89) |
| radiotherapy in last 6/12 | 11890 (0.12) | 1441 (0.11) | 81 (2.09) | 144 (0.77) |
| solid organ transplant<br>ever | 2104 (0.02) | 358 (0.03) | 10 (0.26) | 62 (0.33) |
| COPD | 209519 (2.20) | 17070 (1.32) | 683 (17.61) | 1529 (8.15) |
| Asthma | 1292225 (13.56) | 210812 (16.24) | 567 (14.62) | 3655 (19.49) |
| rare pulmonary<br>conditions | 51203 (0.54) | 5374 (0.41) | 139 (3.58) | 364 (1.94) |
| pulmonary hypertension | 8356 (0.09) | 753 (0.06) | 36 (0.93) | 83 (0.44) |
| coronary heart disease | 325875 (3.42) | 28443 (2.19) | 935 (24.11) | 1962 (10.46) |
| stroke | 201933 (2.12) | 18250 (1.41) | 703 (18.13) | 1432 (7.63) |
| atrial fibrillation | 228054 (2.39) | 20448 (1.58) | 879 (22.67) | 1551 (8.27) |
| congestive cardiac failure | 119691 (1.26) | 10588 (0.82) | 628 (16.19) | 1096 (5.84) |
| VTE | 183548 (1.93) | 20313 (1.57) | 434 (11.19) | 1150 (6.13) |
| peripheral vascular<br>disease | 65488 (0.69) | 4885 (0.38) | 243 (6.27) | 461 (2.46) |
| congenital heart disease | 44255 (0.46) | 7137 (0.55) | 19 (0.49) | 146 (0.78) |
| dementia | 99967 (1.05) | 15272 (1.18) | 1114 (28.73) | 1400 (7.46) |
| parkinson's disease | 22804 (0.24) | 2248 (0.17) | 140 (3.61) | 185 (0.99) |
| epilepsy | 124836 (1.31) | 16878 (1.30) | 116 (2.99) | 537 (2.86) |
| rare neuro conditions | 28958 (0.30) | 4074 (0.31) | 35 (0.90) | 402 (2.14) |
| cerebral palsy | 10915 (0.11) | 1438 (0.11) | * | 43 (0.23) |
| osteoporotic fracture | 373642 (3.92) | 47758 (3.68) | 695 (17.92) | 1429 (7.62) |
| RA or SLE | 238262 (2.50) | 27153 (2.09) | 335 (8.64) | 1206 (6.43) |
| cirrhosis | 21588 (0.23) | 2145 (0.17) | 68 (1.75) | 226 (1.20) |
| bipolar disorder or<br>schizophrenia | 108993 (1.14) | 12527 (0.97) | 75 (1.93) | 427 (2.28) |
| inflammatory bowel<br>disease | 90015 (0.94) | 13643 (1.05) | 70 (1.81) | 519 (2.77) |
| sickle cell disease, HIV or<br>severe combined<br>immunodeficiency | 26579 (0.28) | 3760 (0.29) | 23 (0.59) | 166 (0.89) |

### Factors associated with increased/decreased risk of severe COVID-19 outcomes

Figure 1 shows the adjusted HRs for variables in the final QCOVID4 model for COVID-19 death in men. Figure 2 show the corresponding results for women. Figures 3 and 4 show the adjusted HR for variables in the final model for COVID-19 hospital admission in men and women. The adjusted HRs for fractional polynomial terms for each of the models for age and BMI can be found in supplementary figure 1.

**Figure 1.**
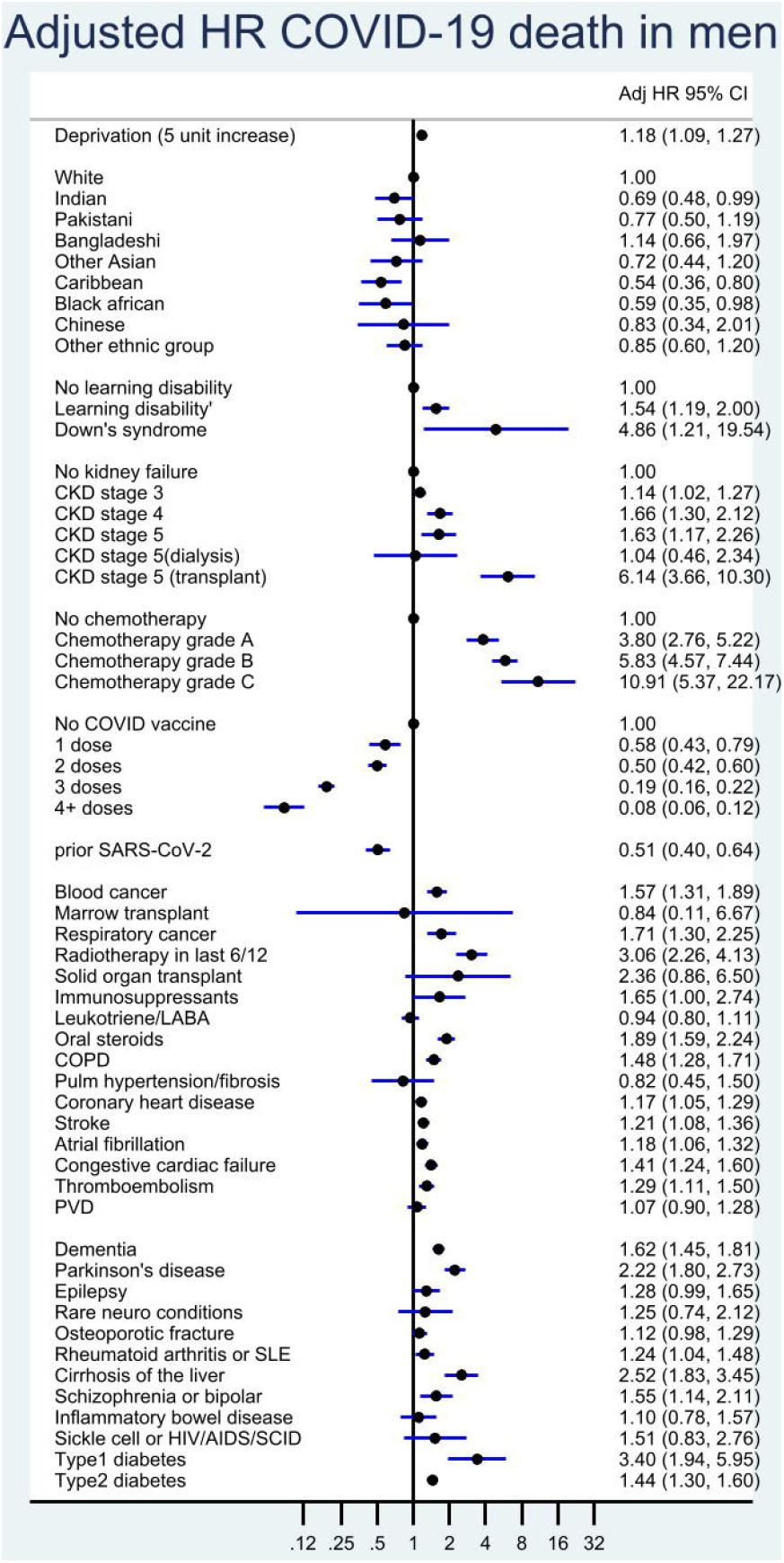
QCOVID4 (mortality): Adjusted hazard ratios for COVID-19 death in men mutually adjusted and also adjusted for fractional polynomial terms for age and BMI.

**Figure 2.**
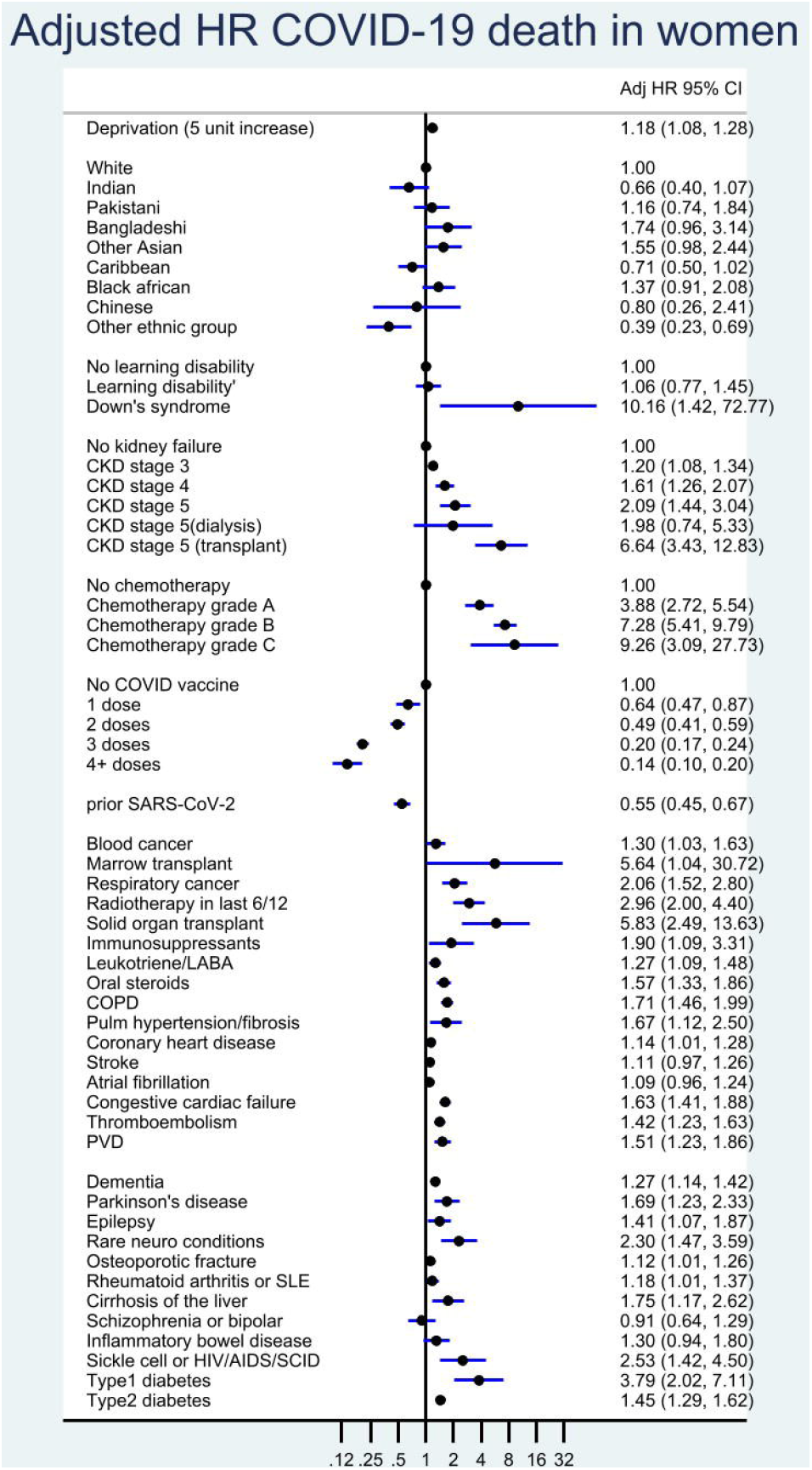
QCOVID4 (mortality): Adjusted hazard ratios for COVID-19 death in women mutually adjusted and also adjusted for fractional polynomial terms for age and BMI.

**Figure 3.**
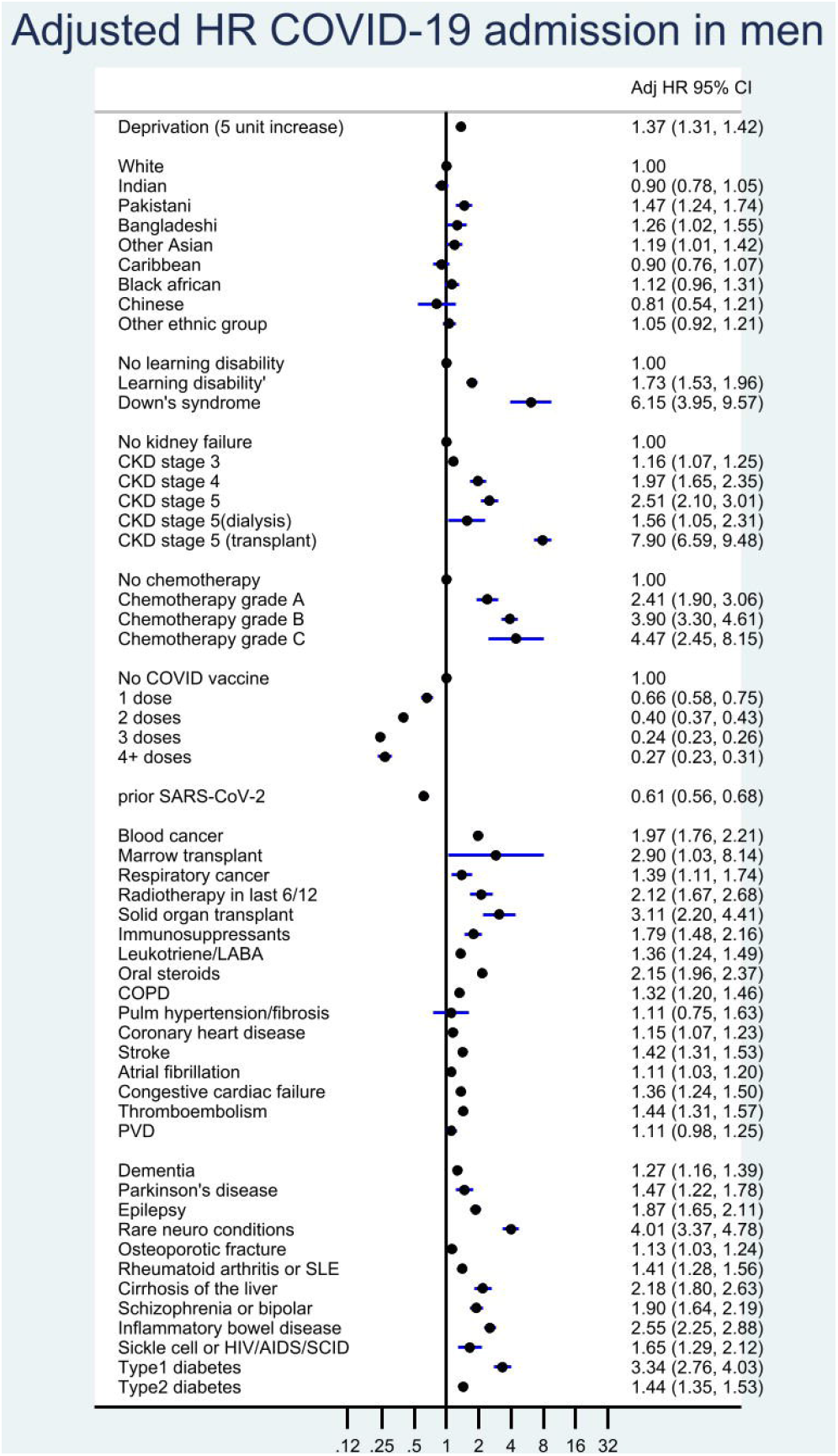
QCOVID4 (admissions): Adjusted hazard ratios for COVID-19 admissions in men mutually adjusted and also adjusted for fractional polynomial terms for age and BMI.

**Figure 4.**
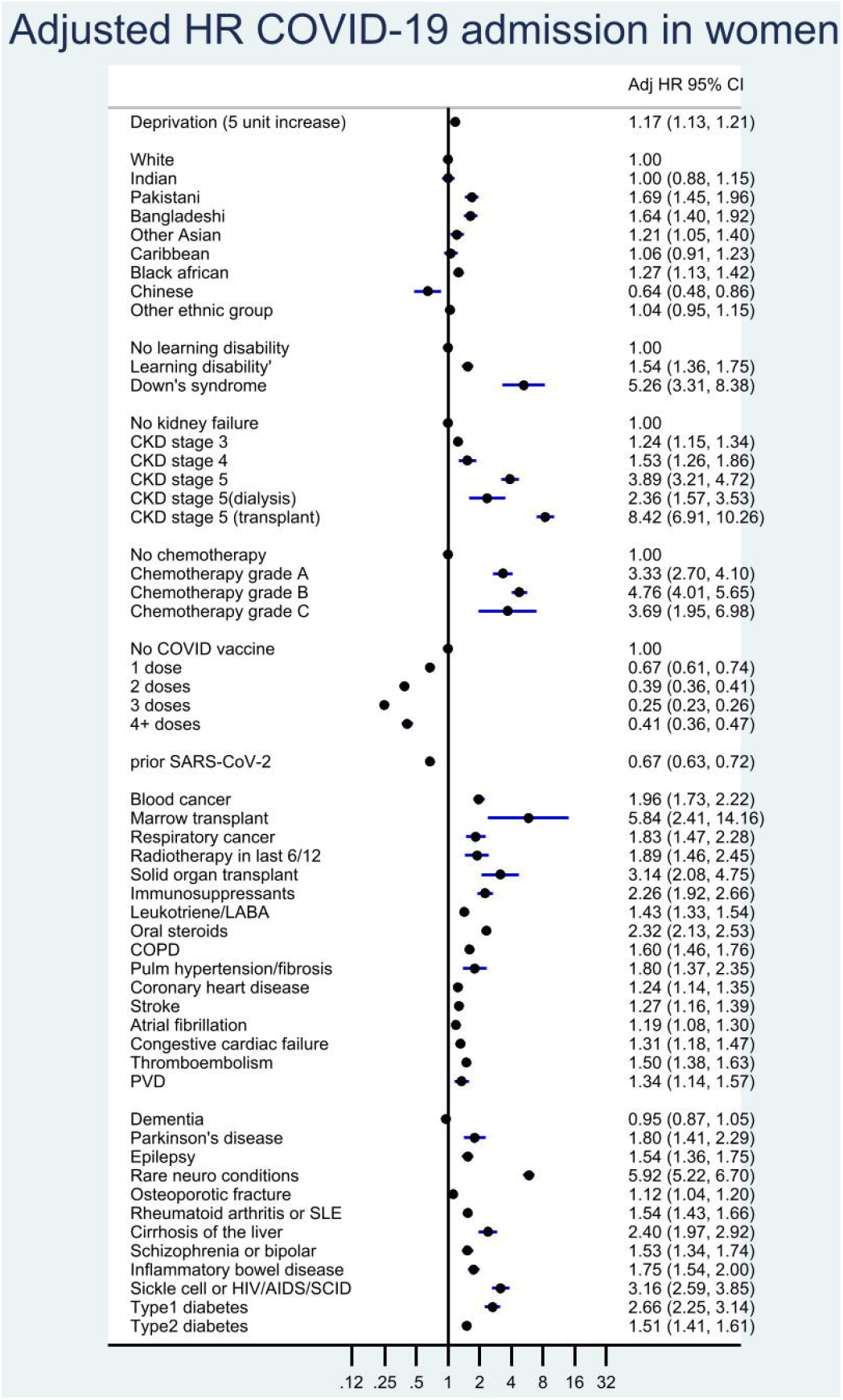
QCOVID4 (admission): Adjusted hazard ratios for COVID-19 admissions in women mutually adjusted and also adjusted for fractional polynomial terms for age and BMI.

The COVID-19 mortality rate in men increased very steeply with age and there was also an association with deprivation. In the final model in men adjusted HRs were highest for those with the following conditions (for 95% CI see Figure 1): kidney transplant (6.1-fold); Down’s syndrome (4.9-fold increase); radiotherapy (3.1-fold); type 1 diabetes (3.4-fold); chemotherapy grade A (3.8-fold), grade B (5.8-fold); grade C (10.9-fold); solid organ transplant ever (2.4-fold); dementia (1.6-fold); Parkinson’s disease (2.2-fold); liver cirrhosis (2.5-fold). Other conditions associated with increased COVID-19 mortality included learning disability, chronic kidney disease (stages 4 and 5), blood cancer, respiratory cancer, immunosuppressants, oral steroids, COPD, coronary heart disease, stroke, atrial fibrillation, heart failure, thromboembolism, rheumatoid/SLE, schizophrenia/bipolar disease sickle cell/HIV/SCID; type 2 diabetes. Unlike QCOVID2, there was no association by residential status or with asthma, rare pulmonary conditions, cerebral palsy or congenital heart disease. There was no difference in risk according to HBAC1C levels, so we included type 1 and type 2 diabetes as binary rather than categorical variables. These results were generally similar in women (Figure 2).

The increased risks of both COVID-19 death and admission in ethnic minority groups observed in previous pandemic waves were either not observed or were much less marked in QCOVID4 compared with earlier analyses. There were no significantly increased risks of COVID-19 death by ethnic group in men or women compared with the white group. There was an increased risk of COVID-19 admission among Bangladeshi, Pakistani and Other Asian men and women and an increased risk of admission for Black African women. There were no other increased risks for Indian, Other Asian, Black ethnicities or Chinese for admission.

The COVID-19 mortality rate was lower among those who had received COVID-19 vaccination compared with unvaccinated individuals with evidence of a dose response relationship as summarised in Supplementary table 2. For example, compared with unvaccinated men, there was a 42% risk reduction associated with one vaccination dose (adjusted HR 0.58 (95% CI 0.43, 0.79)) and a 92% reduction in risk for men with 4 or more doses (adjusted HR 0.08 (95% CI 0.06, 0.12)). The reduced COVID-19 mortality risks associated with COVID-19 vaccination doses were similar in women. COVID-19 admission risk was also reduced in vaccinated men and women.

Prior SARS-CoV-2 infection was associated with a 49% reduced risk of COVID-19 death in men (adjusted HR 0.51 (95% CI 0.40, 0.64)) and a 45% reduced risk in women (adjusted HR 0.55 (95% 0.45 to 0.67)), independent of age, ethnicity, vaccination status and other factors included in the final QCOVID4 models. The 39% reduction in admission risk for men (adjusted HR 0.61 (95% 0.56, 0.68)) was similar to the 33% reduction in admission risk for women (adjusted HR 0.67 (95% 0.63, 0.72)).

### Discrimination

Table 2 shows the explained variation and discrimination of QCOVID1, QCOVID2 and QCOVID4 models in the validation cohort for women and men overall for COVID-19 death and hospital admission. The QCOVID4 algorithm explained 76.6% (95%CI 74.4 to 78.8) of the variation (R^2^) in time to COVID-19 death in women. The D statistic was 3.70 (95%CI 3.48 to 3.93) and the Harrell’s C statistic was 0.965 (95%CI 0.951 to 0.978). The corresponding results for COVID-19 death in men were similar with R of 76.0% (95% 73.9 to 78.2); D statistic 3.65 (95%CI 3.43 to 3.86) and a C statistic of 0.970 (95%CI 0.962 to 0.979).

**TABLE 2:** Performance of the QCOVID1, 2 and 4 algorithms in males and females in validation cohort.

| statistic | *COVID-19 death* |  | *COVID-19 admission* |  |
| --- | --- | --- | --- | --- |
|  | females | males | females | males |
| **QCOVID1** |  |  |  |  |
| R<sup>2</sup> | 73.60 (71 to 76.1) | 73.0 (70.6 to 75.4) | 27.6 (24.7 to 30.5) | 48.4 (45.6 to 51.2) |
| D statistic | 3.41 (3.19 to 3.64) | 3.36 (3.16 to 3.57) | 1.26 (1.17 to 1.36) | 1.98 (1.87 to 2.09) |
| Harrell's C | .945 (.928 to .963) | .948 (.933 to .963) | .684 (.669 to .7) | .798 (.781 to .814) |
| **QCOVID2** |  |  |  |  |
| R<sup>2</sup> | 75.4 (73.1 to 77.7) | 74.1 (71.8 to 76.4) | 67.4 (64.1 to 70.7) | 68.2 (65.2 to 71.2) |
| D statistic | 3.58 (3.36 to 3.81) | 3.46 (3.25 to 3.67) | 2.94 (2.72 to 3.16) | 3.00 (2.79 to 3.21) |
| Harrell's C | .968 (.959 to .978) | .959 (.946 to .971) | .939 (.93 to .948) | .932 (.918 to .945) |
| **QCOVID4** |  |  |  |  |
| R<sup>2</sup> | 76.6 (74.4 to 78.8) | 76 (73.9 to 78.2) | 73.9 (71.5 to 76.3) | 74.4 (72.1 to 76.6) |
| D statistic | 3.70 (3.48 to 3.93) | 3.65 (3.43 to 3.86) | 3.44 (3.23 to 3.66) | 3.49 (3.28 to 3.69) |
| Harrell's C | .965 (.951 to .978) | .97 (.962 to .979) | .965 (.956 to .973) | .97 (.963 to .977) |

The performance of QCOVID4 for COVID-19 mortality was slightly improved compared with both QCOVID1 and QCOVID2. For COVID-19 hospital admissions, however, QCOVID4 had significantly improved performance compared with QCOVID2, which in turn had improved performance compared with QCOVID1. For example, Harrell’s C statistic in men for QCOVID4 was 0.970 (95%CI 0.963 to 0.977) compared with 0.932 (95%CI 0.918 to 0.945) for QCOVID2 and 0.798 (95%CI 0.781 to 0.814), for QCOVID1.

Supplementary table 3 shows performance of QCOVID4 in subgroups by age and ethnicity. Performance measures were generally higher in the younger age groups and similar across ethnic groups (where there were sufficient numbers in the subgroup to enable an analysis).

### Calibration

Figures 5 and 6 show the mean predicted risks and the observed risks for COVID-19 mortality and admission using QCOVID4 to assess calibration in the validation cohort. There was close correspondence between the mean predicted risks and the observed risks within each model twentieth in women and men indicating the algorithms are well calibrated.

**Figure 5.**
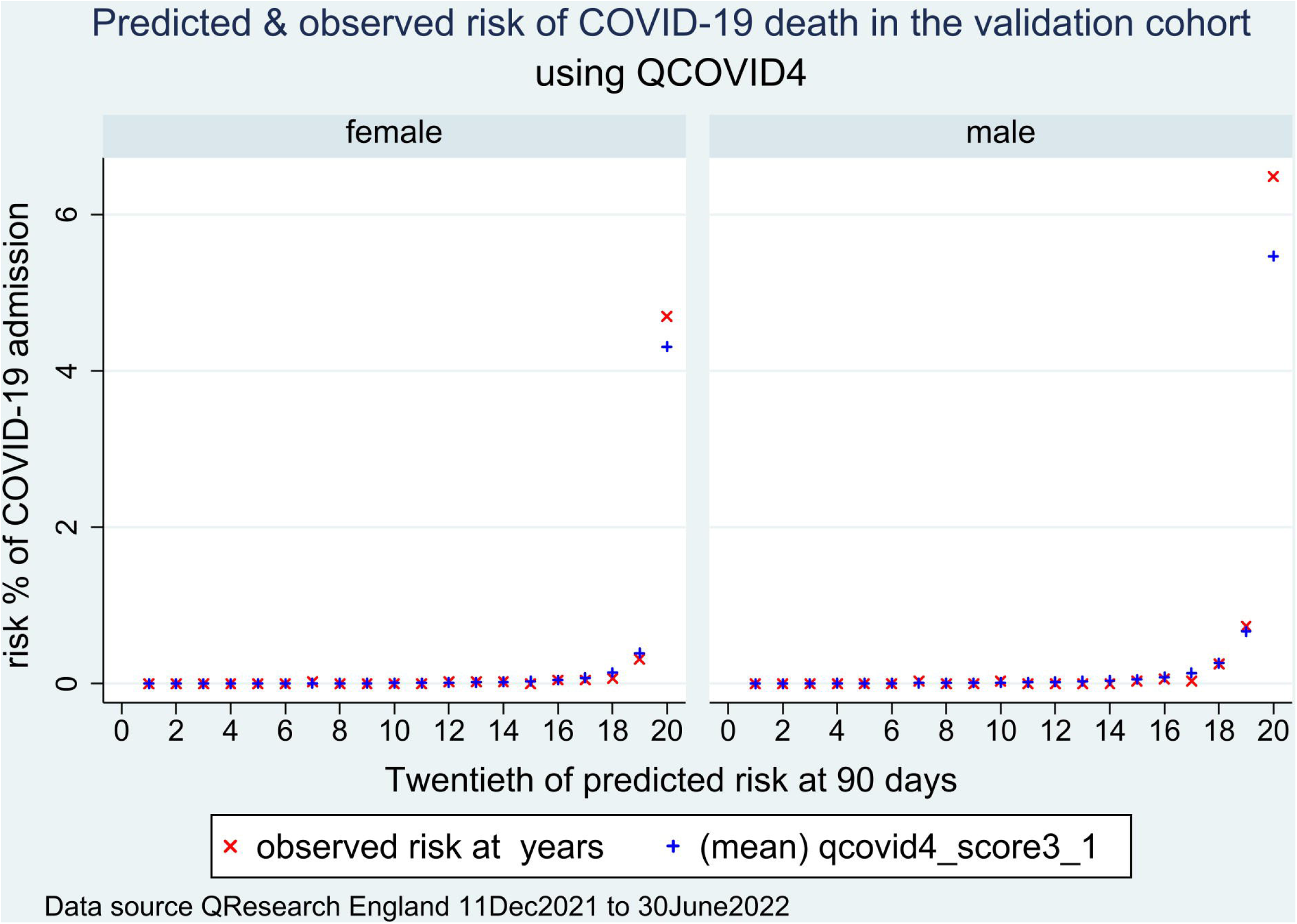
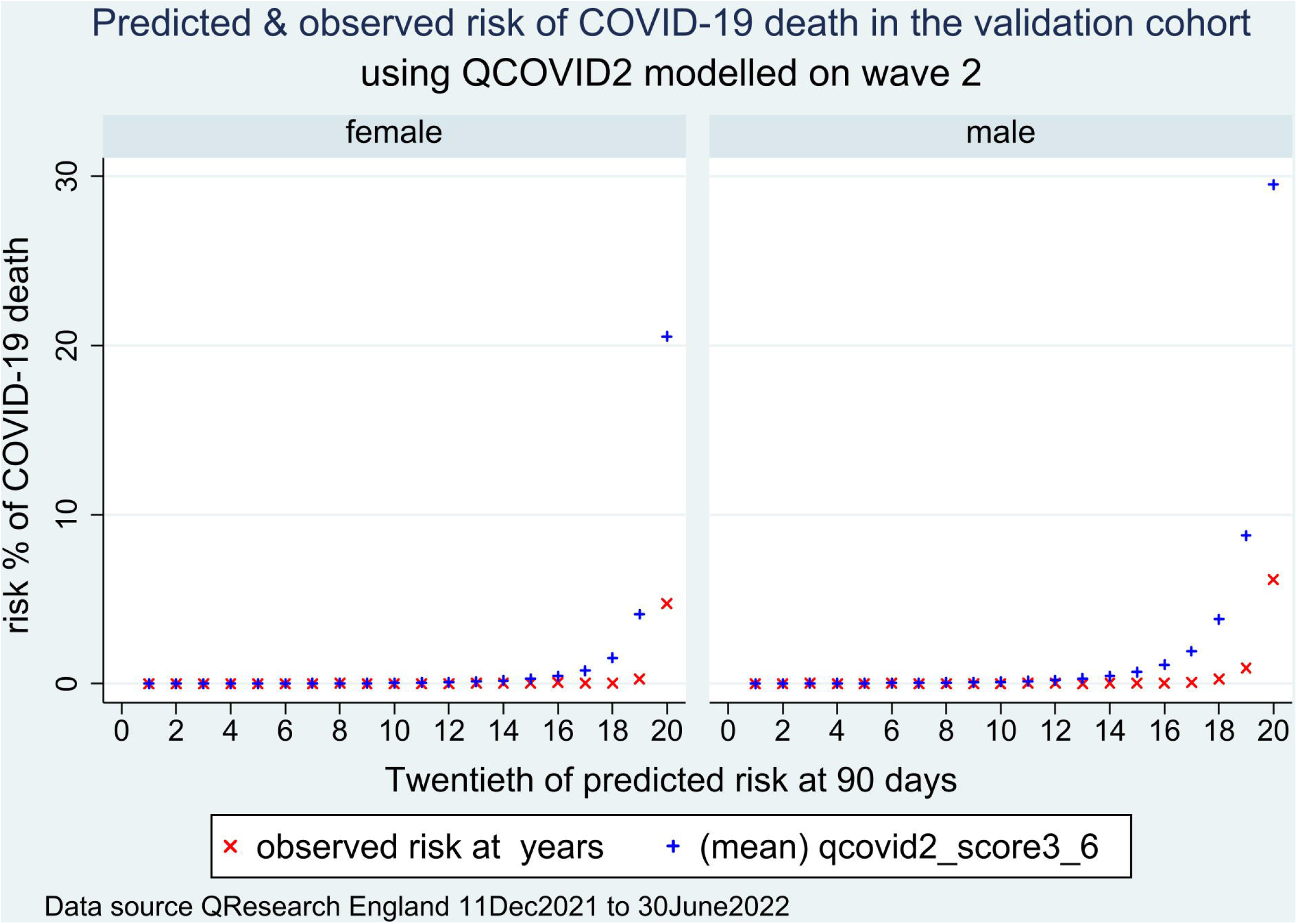
Calibration of the QCOVID4 risk model to predict COVID-19 death following vaccination.

**Figure 6.**
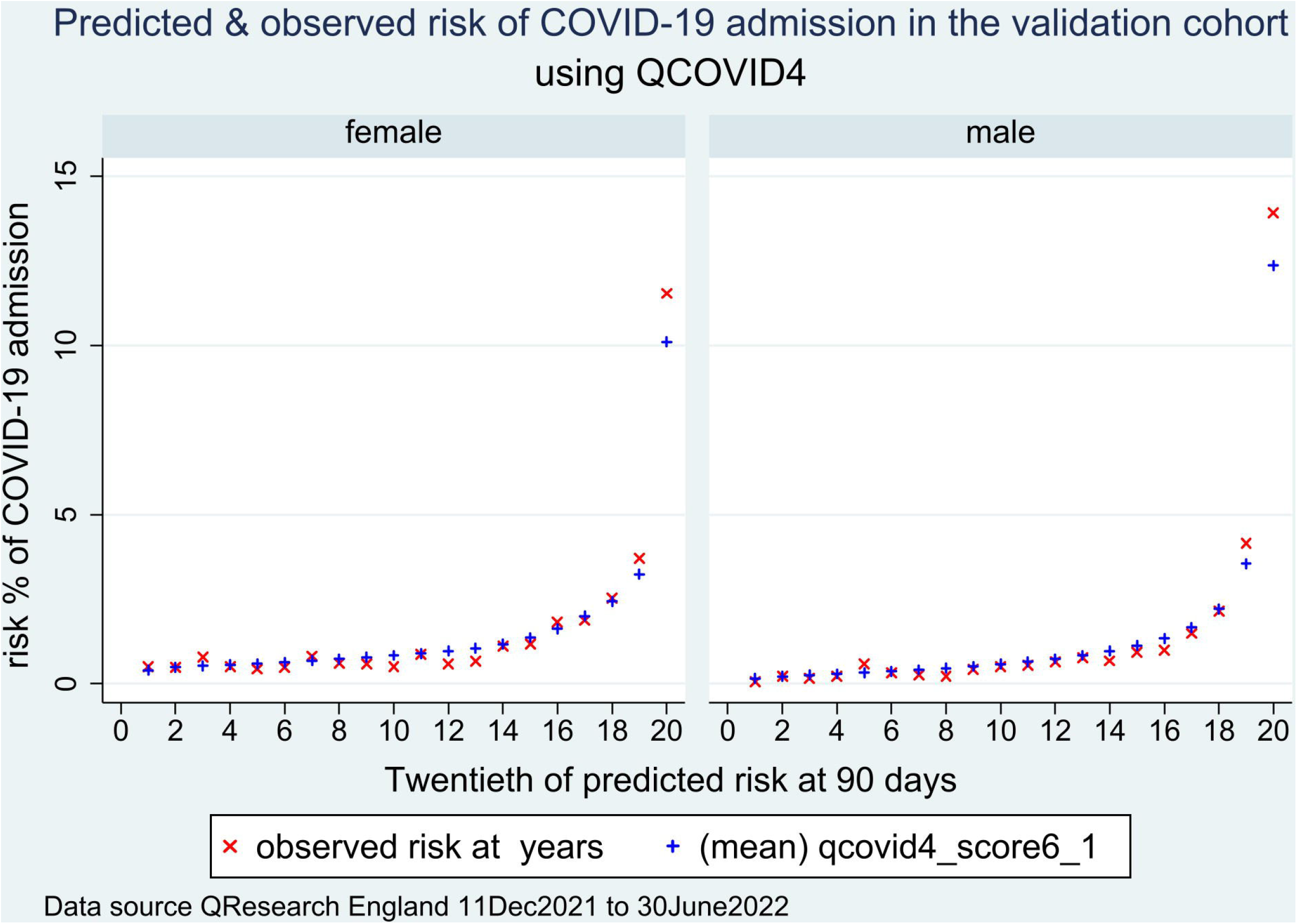
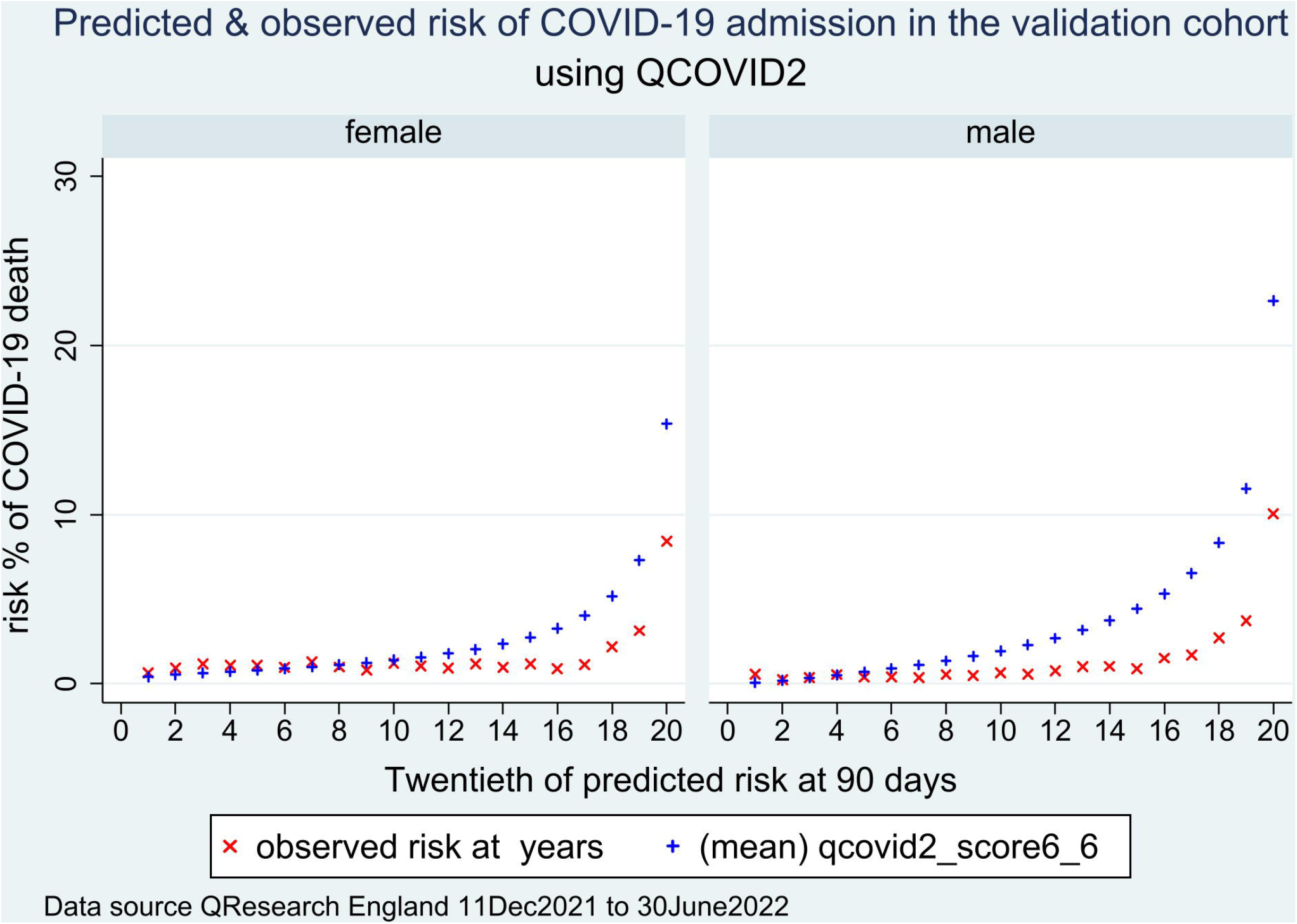
Calibration of the QCOVID4 risk model to predict COVID-19 admission following vaccination.

Supplementary figures 2 and 3 show the corresponding results for QCOVID2 which has a degree of miscalibration indicating over prediction.

### Thresholds

Table 3 shows the classification statistics in the validation cohort for men and women by twentieths of predicted mortality risk using QCOVID4. For example, for the 20% of the cohort at highest predicted risk (i.e. those with a 90-day predicted risk score of 0.075% or higher), the sensitivity was 97.8%, specificity was 80.2% and the observed risk was 1.54%. The corresponding figures for the top 5% at highest predicted risk were a sensitivity of 87.6%, specificity of 95.3% and observed 90-day risk of 5.52%.

**TABLE 3.** sensitivity, specificity and observed 90-day morality risk at different centiles of predict risk using QCOVID4 to predict COVID-19 mortality (461 deaths) in the validation cohort of 145,397 people with a SARS-CoV-2 positive test result.

| *Top centile* | *Predicted 90 day risk Threshold (%)* | *sensitivity* | *specificity* | *Observed risk at 90 days (%)* |
| --- | --- | --- | --- | --- |
| top 5% | 0.908 | 87.6 | 95.3 | 5.520 (5.018, 6.070) |
| top 10% | 0.278 | 94.8 | 90.3 | 2.986 (2.722, 3.276) |
| top 15% | 0.129 | 96.7 | 85.3 | 2.032 (1.853, 2.228) |
| top 20% | 0.075 | 97.8 | 80.2 | 1.541 (1.406, 1.689) |
| top 25% | 0.049 | 98.5 | 75.2 | 1.241 (1.132, 1.360) |
| top 30% | 0.034 | 98.5 | 70.2 | 1.034 (0.943, 1.133) |
| top 35% | 0.025 | 98.5 | 65.2 | 0.886 (0.809, 0.972) |
| top 40% | 0.019 | 98.7 | 60.2 | 0.777 (0.709, 0.852) |
| top 45% | 0.015 | 99.1 | 55.2 | 0.694 (0.633, 0.761) |
| top 50% | 0.012 | 99.3 | 50.2 | 0.626 (0.571, 0.686) |
| top 55% | 0.010 | 99.3 | 45.1 | 0.569 (0.519, 0.624) |
| top 60% | 0.008 | 99.6 | 40.1 | 0.523 (0.477, 0.573) |
| top 65% | 0.007 | 99.6 | 35.1 | 0.483 (0.440, 0.529) |
| top 70% | 0.006 | 99.6 | 30.1 | 0.448 (0.409, 0.491) |
| top 75% | 0.005 | 99.8 | 25.1 | 0.419 (0.382, 0.459) |
| top 80% | 0.004 | 100.0 | 20.1 | 0.394 (0.359, 0.431) |
| top 85% | 0.003 | 100.0 | 15.0 | 0.371 (0.338, 0.406) |
| top 90% | 0.0025 | 100.0 | 10.0 | 0.350 (0.319, 0.384) |
| top 100% | 0.0017 | 100.0 | 5.0 | 0.332 (0.303, 0.363) |

We identified 34,864 patients in the NHS Digital high-risk cohort in the QResearch cohorts of whom 3,600 were in the validation cohort. Supplementary Table 4 shows the characteristics of these patients compared with the characteristics of 3,600 patients (top 2.48%) with the highest predicted risks of COVID-19 mortality using the QCOVID4 models. Patients in the QCOVID4 high risk group tended to be much older (mean 85.0 years compared with 55.4 years) with higher levels of co-morbidities, with some exceptions (e.g. CKD5, blood cancer, grade B chemotherapy, rare neurological conditions). There were 520 patients included in both high-risk groups. Of the 461 COVID19 deaths which occurred in the validation cohort, 333 (72.2%) occurred in the QCOVID4 high risk group and 95 (20.6%) in the NHS Digital high-risk group. Uptake of the COVID-19 therapeutics (both antivirals and nMABS) was very low with 504 (14.0%) of the NHS digital high-risk group receiving treatment and 131 (3.6%) of the QCovid4 high risk cohort.

## DISCUSSION

### Principal findings

We have developed and validated a new QCOVID model (QCOVID4) using data recorded during the Omicron wave in England. QCOVID4 more accurately identifies individuals at highest levels of absolute risk for targeted interventions than the ‘conditions-based’ approach adopted by NHS Digital based on relative risk of a list of medical conditions.

We also compared performance with earlier versions of the QCOVID algorithms on this dataset. The earlier QCOVID models were developed in the first wave of the original variant (QCOVID1) and second waves of Alpha and Delta variants (QCOVID2). Overall, the factors associated with increased risk in earlier models ^1 6^, were still associated with increased risk in the QCOVID4 model. An exception was ethnic minority groups where the previously elevated risks, particularly associated with South Asian and Black ethnicities for COVID-19 death in QCOVID1^1^ and QCOVID2 ^6^, were no longer apparent in QCOVID4. There was however, a residual increased risk for Pakistani and Bangladeshi groups for COVID-19 admission compared with the White group for both men and women, and an increased admission risk for Black African and other Asian women after adjustment for age, ethnicity, deprivation, co-morbidity and vaccination status.

We have demonstrated that infection with SARS-CoV-2 prior to the study period was associated with approximately 50% lower risk of COVID-19 mortality in both men and women. This was independent of age, ethnicity, deprivation, co-morbidity and vaccination status. Similarly, there was a dose-dependent reduction in mortality risk in men and women following COVID-19 vaccination with each subsequent dose conferring additional benefits.

The validation shows that all three models (QCOVID4, QCOVID1 and QCOVID2) have high levels of discrimination for COVID-19 mortality and explained variation in this dataset. The QCOVID4 model has substantially improved discrimination and explained variation for predicting risk of COVID-19 hospital admission. Of those identified by NHS Digital as a high risk group for targeted therapeutics (2.5% of total), only 14% were also identified in an equivalently sized high-risk group using QCOVID4. Nearly three quarters of the total COVID-19 deaths in the validation cohort occurred in the QCOVID4 high risk group compared with one fifth in the NHS Digital cohort. This difference was not explained by the use of therapeutic interventions which was low in both groups.

The validation results also demonstrate that the QCOVID4 model is well-calibrated to the current contemporaneous validation dataset. QCOVID1 was developed in a general population to estimate the combined risk of ‘catching and dying’ from COVID-19 due to lack of testing data available in the first pandemic and so estimates of absolute risk would not be valid for prediction of COVID-19 death in people with a SARS-CoV-2 positive test. There was over-prediction associated with QCOVID2 which is likely to mainly reflect higher levels of vaccination, better treatments and differences in the variant type with Omicron now generally considered to be less severe than earlier variants ^7 27^. Taken together, QCOVID1 and QCOVID2 have acceptable ongoing utility for ranking those at highest risk of death for interventions but are less robust for predicting the absolute of risk of each outcome than QCOVID4.

### Strengths and limitations of this study

Our study has some major strengths, but some important limitations. These include specific issues related to COVID-19 along with others similar to those for a range of other widely used clinical risk prediction algorithms developed using the QResearch database ^28 29 30^. Key strengths include the use of very large representative population-based contemporaneous data sources which have been used to develop other widely used risk prediction tools ^28 29^, the wealth of candidate risk predictors, the prospective recording of outcomes and their ascertainment using multiple national-level database linkage, lack of selection, recall and respondent biases, and robust statistical analysis. We have used non-linear terms to model association with BMI and age and multiple imputation to handle missing data.

Limitations include relatively small numbers of events in some of the subgroups which is an inevitable consequence of undertaking an analysis involving multiple subgroups during a relatively short pandemic wave. Whilst we have accounted for many risk factors for severe COVID-19 outcomes, there may be risks conferred by some rare medical conditions or other factors associated with exposure such as occupation that are poorly recorded in general practice or hospital records and which may be being proxied to some extent by the covariates included. Also our study does not address outcomes related to the very recent Omicron BA.4/BA.5 wave in England which was identified as a variant of concern on 18^th^ May 2022.

Whilst we have reported a validation using practices from QResearch, these practices were completely separate to those used to develop the model. Previously we have used this approach to develop and validate other widely used prediction models. When these models have been validated on data from different clinical computer systems, the results have been very similar ^31-33^. Work has now been completed to evaluate earlier QCOVID models in external datasets including the English national dataset hosted by the Office of National Statistics ^2^, Scotland^4^ and Wales^3^ data including data which has not been used to derive the algorithm and these evaluations also showed similar levels of performance to the validation in QResearch practices.

### Implications for clinical practice, policy and research

The utility of a risk prediction model crucially depends on the purpose for which it has been designed and the setting in which it has been developed and that where it might be deployed. The speed at which new COVID-19 variants of concern have emerged and become dominant inevitably means that prediction models could become out of date almost as soon as they have been developed and implemented. This study, with its comparisons across the last three major pandemic waves, each associated with different variant types, provides evidence that the performance of QCOVID1 and QCOVID2 algorithms remains good and therefore are likely to be effective for risk stratifying or ranking individuals for interventions. Algorithms may need recalibration before being used to calculate absolute risks in settings or time periods with different mortality or admission rates.

Although large disparities among ethnic minority groups were observed during the early waves of the pandemic, there were no increased mortality risks for any ethnic group. We observed increased risks of admission for Bangladeshi, Pakistani and Other Asian men and women and for Black African women. There were no other increased risks for Indian, Other Asian, Black ethnicities or Chinese for admission. Reports from the second wave showed higher rates of COVID-19 mortality among Pakistani and Bangladeshi groups^34^ suggesting ethnic disparities may have been improved by widespread vaccination and other public health interventions to reduce risk of exposure or infection.

## Conclusion

In conclusion, the QCOVID4 algorithm developed using data from the Omicron wave has excellent performance for identifying those at highest risk of severe COVID-19 outcomes. This can be used to risk stratify patients for intervention (such as COVID-19 therapeutics) and inform clinical decision making on individualised risk management with patients and this could be more effective than an approach based on relative risks of individual medical conditions.

## Data Availability

To guarantee the confidentiality of personal and health information only the authors have had access to the data during the study in accordance with the relevant licence agreements. Access to the QResearch data is according to the information on the QResearch website (www.qresearch.org). The full model, model coefficients, functional form and cumulative incidence function, is published on the qcovid.org website.

https://www.qcovid.org

## CONTRIBUTORS

Study conceptualisation was led by JHC and CC. JHC specified the data, organised data approvals and data linkage. JHC and CC, designed the statistical analysis plan. JHC and CC undertook the analyses. JHC wrote the first draft of the paper. AS, KK and JVT contributed to development of study ideas, analysis plan and provided critical comments on the manuscript. The Office of the Chief Medical Officer contributed to the development of the study question and facilitated access to relevant national datasets, contributed to interpretation of data, drafting of the report. All authors contributed to the interpretation of the results and revision of the manuscript and approved the final version of the manuscript.

## FUNDING

This study is funded by the National Institute for Health Research (NIHR) following a commission by Department of Health and Social Care. The researchers are independent from the NIHR. The QResearch is supported by funds from the John Fell Oxford University Press Research Fund, grants from Cancer Research UK (CR-UK) grant number C5255/A18085, through the Cancer Research UK Oxford Centre, grants from the Oxford Wellcome Institutional Strategic Support Fund (204826/Z/16/Z), during the conduct of the study.

## COMPETING INTERESTS

All authors have completed the ICMJE uniform disclosure form at www.icmje.org/coi_disclosure.pdf and declare: JHC reports grants from National Institute for Health Research (NIHR) Biomedical Research Centre, Oxford, grants from John Fell Oxford University Press Research Fund, grants from Cancer Research UK (CR-UK) grant number C5255/A18085, through the Cancer Research UK Oxford Centre, grants from the Oxford Wellcome Institutional Strategic Support Fund (204826/Z/16/Z) and other research councils, during the conduct of the study. JHC is an unpaid director of QResearch, a not-for-profit organisation which is a partnership between the University of Oxford and EMIS Health who supply the QResearch database used for this work. JHC is a founder and shareholder of ClinRisk ltd and was its medical director until 31^st^ May 2019. ClinRisk Ltd produces open and closed source software to implement clinical risk algorithms (outside this work) into clinical computer systems. JHC is chair of the NERVTAG risk stratification subgroup and a member of SAGE COVID-19 groups and the NHS group advising on prioritisation of use of monoclonal antibodies in COVID-19 infection. CC reports receiving personal fees from ClinRisk Ltd, outside this work and is a member of the NERVTAG risk stratification subgroup. KK is Chair of the Ethnic Subgroup of the UK Scientific Advisory Group for Emergencies (SAGE) and a member of SAGE. AS has received research grants from NIHR, MRC, HDRUK, CSO, NCS, GSK for COVID-19 work and has provided advice for AstraZeneca’s Thrombotic Thrombocytopenic Taskforce and the UK and Scottish Government COVID-19 Advisory Groups (roles unremunerated)

## ETHICS APPROVAL

The QResearch^®^ ethics approval was provided on 8^th^ June 2020 by the East Midlands-Derby Research Ethics Committee [reference 18/EM/0400].

## DATA SHARING

To guarantee the confidentiality of personal and health information only the authors have had access to the data during the study in accordance with the relevant licence agreements. Access to the QResearch data is according to the information on the QResearch website (www.qresearch.org). The full model, model coefficients, functional form and cumulative incidence function, is published on the qcovid.org website. https://www.qcovid.org/Home/Algorithm.

## TRANSPARENCY

JHC had full access to all data in the study and takes responsibility for the integrity of the data and the accuracy of the data analysis. JHC is the guarantor for the study and affirms that the manuscript is an honest, accurate, and transparent account of the study reported; that no important aspects of the study have been omitted and that any discrepancies from the study as planned have been explained.

## ACKNOWLEDGMENTS

We acknowledge the contribution of EMIS practices who contribute to QResearch^®^ and EMIS Health and the Universities of Nottingham and Oxford for expertise in establishing, developing or supporting the QResearch database. This project involves data derived from patient-level information collected by the NHS, as part of the care and support of cancer patients. The data is collated, maintained and quality assured by the National Cancer Registration and Analysis Service, which is now part NHS Digital. The Hospital Episode Statistics data, SARS-Cov-2 results and civil registration mortality data are used by permission from NHS Digital who retain the copyright in that data. NHS Digital and Public Health England bears no responsibility for the analysis or interpretation of the data. KK is supported by NIHR Applied Research Collaboration East Midlands (ARC-EM) and the NIHR BRC.

## COPYRIGHT STATEMENT

“I, the Submitting Author has the right to grant and does grant on behalf of all authors of the Work (as defined in the *<u>author licence</u>*), an exclusive licence on a worldwide, perpetual, irrevocable, royalty-free basis to BMJ Publishing Group Ltd (“BMJ”) its licensees. The Submitting Author accepts and understands that any supply made under these terms is made by BMJ to the Submitting Author unless I am acting as an employee on behalf of my employer which is paying any applicable article publishing charge (“APC”) for Open Access articles. Where the Submitting Author wishes to make the Work available on an Open Access basis (and intends to pay the relevant APC), the terms of reuse of such Open Access shall be governed by a Creative Commons licence – details of these licences and which licence will apply to this Work are set out in our licence referred to above.”

**SUPPLEMENTARY TABLE 1.** Baseline characteristics of the validation cohort of patients with a SARS-CoV-2 positive test, COVID-19 death and COVID-19 admission.

|  | *total population* | *SARS-CoV-2*<br>*positive* | *COVID-19 death* | *COVID-19*<br>*admission* |
| --- | --- | --- | --- | --- |
| total | 1,064,255 | 145,397 | 461 | 2,124 |
| males (%) | 531631 (49.95) | 62305 (42.85) | 240 (52.06) | 846 (39.83) |
| mean age (SD) | 47.85 (18.49) | 42.96 (16.41) | 81.30 (11.87) | 55.79 (22.10) |
| White | 699679 (65.74) | 101478 (69.79) | 356 (77.22) | 1474 (69.40) |
| Indian | 23101 (2.17) | 2897 (1.99) | * | 33 (1.55) |
| Pakistani | 26677 (2.51) | 2291 (1.58) | * | 59 (2.78) |
| Bangladeshi | 16650 (1.56) | 1550 (1.07) | * | 29 (1.37) |
| Other Asian | 16571 (1.56) | 2046 (1.41) | * | 26 (1.22) |
| Caribbean | 8336 (0.78) | 959 (0.66) | 5 (1.08) | 33 (1.55) |
| Black african | 26218 (2.46) | 2743 (1.89) | * | 56 (2.64) |
| Chinese | 9650 (0.91) | 902 (0.62) | * | 11 (0.52) |
| Other ethnic group | 39788 (3.74) | 4931 (3.39) | 6 (1.30) | 77 (3.63) |
| Ethnicity not recorded | 197585 (18.57) | 25600 (17.61) | 80 (17.35) | 326 (15.35) |
| 1 (most affluent) | 273276 (25.68) | 38571 (26.53) | 125 (27.11) | 478 (22.50) |
| 2 | 239009 (22.46) | 34309 (23.60) | 128 (27.77) | 420 (19.77) |
| 3 | 199585 (18.75) | 28114 (19.34) | 97 (21.04) | 417 (19.63) |
| 4 | 175259 (16.47) | 22214 (15.28) | 62 (13.45) | 389 (18.31) |
| 5 (most deprived) | 163433 (15.36) | 19639 (13.51) | 48 (10.41) | 382 (17.98) |
| 6 (not recorded) | 13693 (1.29) | 2550 (1.75) | * | 38 (1.79) |
| neither | 1054342 (99.07) | 143343 (98.59) | 350 (75.92) | 1989 (93.64) |
| Care home | 8137 (0.76) | 1903 (1.31) | 110 (23.86) | 125 (5.89) |
| Homeless | 1776 (0.17) | 151 (0.10) | * | 10 (0.47) |
| BMI < 18.5 | 24788 (2.33) | 3443 (2.37) | 37 (8.03) | 69 (3.25) |
| BMI 18.5-24.99 | 302741 (28.45) | 43886 (30.18) | 140 (30.37) | 571 (26.88) |
| BMI 25-29.99 | 260970 (24.52) | 35152 (24.18) | 108 (23.43) | 517 (24.34) |
| BMI 30-34.99 | 123125 (11.57) | 16520 (11.36) | 57 (12.36) | 324 (15.25) |
| BMI 35+ | 47646 (4.48) | 6825 (4.69) | 22 (4.77) | 128 (6.03) |
| BMI 40+ | 24866 (2.34) | 3655 (2.51) | 11 (2.39) | 51 (2.40) |
| BMI not recorded | 280119 (26.32) | 35916 (24.70) | 86 (18.66) | 464 (21.85) |
| no CKD | 1020120 (95.85) | 141747 (97.49) | 293 (63.56) | 1830 (86.16) |
| CKD5 only | 1226 (0.12) | 164 (0.11) | 7 (1.52) | 28 (1.32) |
| CKD5 with dialysis | 369 (0.03) | 53 (0.04) | * | * |
| CKD5 with transplant | 592 (0.06) | 113 (0.08) | * | 27 (1.27) |
| no learning disability | 1044039 (98.10) | 142758 (98.18) | 454 (98.48) | 2059 (96.94) |
| Learning disability | 19731 (1.85) | 2553 (1.76) | 6 (1.30) | 62 (2.92) |
| Downs | 485 (0.05) | 86 (0.06) | * | * |
| No chemo in last 12 | 1060048 (99.60) | 144888 (99.65) | 432 (93.71) | 2065 (97.22) |
| Chemo group A | 1945 (0.18) | 207 (0.14) | 9 (1.95) | 15 (0.71) |
| chemo group B | 2154 (0.20) | 284 (0.20) | 19 (4.12) | 42 (1.98) |
| chemo group C | 108 (0.01) | 18 (0.01) | * | * |
| no type1 diabetes | 1059034 (99.51) | 144553 (99.42) | 459 (99.57) | 2094 (98.59) |
| type 1 HBA<=59 | 1515 (0.14) | 275 (0.19) | * | 9 (0.42) |
| type1 HBA1C 59+ | 3056 (0.29) | 463 (0.32) | * | 18 (0.85) |
| type 1 HBA1C no record | 650 (0.06) | 106 (0.07) | * | * |
| no type2 diabetes | 992924 (93.30) | 139075 (95.65) | 340 (73.75) | 1839 (86.58) |
| type 2 HBA<=59 | 40138 (3.77) | 3620 (2.49) | 76 (16.49) | 152 (7.16) |
| type2 HBA1C 59+ | 22464 (2.11) | 1932 (1.33) | 33 (7.16) | 99 (4.66) |
| type 2 HBA1C no record | 8729 (0.82) | 770 (0.53) | 12 (2.60) | 34 (1.60) |
| no covid vaccine doses | 181718 (17.07) | 15725 (10.82) | 36 (7.81) | 416 (19.59) |
| 1 dose | 31020 (2.91) | 4985 (3.43) | 12 (2.60) | 89 (4.19) |
| 2 doses | 169936 (15.97) | 39637 (27.26) | 87 (18.87) | 418 (19.68) |
| 3 doses | 587386 (55.19) | 84272 (57.96) | 317 (68.76) | 1136 (53.48) |
| 4 doses | 94195 (8.85) | 778 (0.54) | 9 (1.95) | 65 (3.06) |
| SARS-CoV-2 infection prior to study | 147803 (13.89) | 16014 (11.01) | 27 (5.86) | 134 (6.31) |
| Blood cancer | 8393 (0.79) | 998 (0.69) | 38 (8.24) | 91 (4.28) |
| Bone marrow transplant in previous 6 months | 38 (0.00) | * | * | * |
| Respiratory cancer | 2258 (0.21) | 211 (0.15) | 14 (3.04) | 16 (0.75) |
| radiotherapy in last 6/12 | 1401 (0.13) | 181 (0.12) | 8 (1.74) | 25 (1.18) |
| solid organ transplant | 233 (0.02) | 49 (0.03) | 1 (0.22) | 9 (0.42) |
| COPD | 23367 (2.20) | 1889 (1.30) | 72 (15.62) | 180 (8.47) |
| Asthma | 146396 (13.76) | 23767 (16.35) | 76 (16.49) | 378 (17.80) |
| rare pulmonary conditions | 6016 (0.57) | 601 (0.41) | 17 (3.69) | 42 (1.98) |
| pulmonary hypertension | 937 (0.09) | 87 (0.06) | 7 (1.52) | 6 (0.28) |
| coronary heart disease | 37471 (3.52) | 3312 (2.28) | 104 (22.56) | 230 (10.83) |
| stroke | 23218 (2.18) | 2020 (1.39) | 89 (19.31) | 141 (6.64) |
| atrial fibrillation | 26343 (2.48) | 2457 (1.69) | 135 (29.28) | 163 (7.67) |
| congestive cardiac failure | 13976 (1.31) | 1251 (0.86) | 69 (14.97) | 109 (5.13) |
| VTE | 20761 (1.95) | 2269 (1.56) | 46 (9.98) | 127 (5.98) |
| PVD | 7500 (0.70) | 586 (0.40) | 32 (6.94) | 47 (2.21) |
| congenital heart disease | 5234 (0.49) | 821 (0.56) | * | 13 (0.61) |
| dementia | 11518 (1.08) | 1630 (1.12) | 119 (25.81) | 160 (7.53) |
| Parkinson's disease | 2550 (0.24) | 246 (0.17) | 23 (4.99) | 24 (1.13) |
| epilepsy | 14250 (1.34) | 1911 (1.31) | 12 (2.60) | 64 (3.01) |
| rare neurological conditions | 3344 (0.31) | 465 (0.32) | 7 (1.52) | 47 (2.21) |
| cerebral palsy | 1352 (0.13) | 163 (0.11) | * | 7 (0.33) |
| osteoporotic fracture | 43404 (4.08) | 5429 (3.73) | 85 (18.44) | 177 (8.33) |
| RA or SLE | 28446 (2.67) | 3310 (2.28) | 43 (9.33) | 153 (7.20) |
| cirrhosis | 2413 (0.23) | 241 (0.17) | 6 (1.30) | 23 (1.08) |
| bipolar disorder or<br>schizophrenia | 12029 (1.13) | 1392 (0.96) | * | 47 (2.21) |
| IBS | 10167 (0.96) | 1600 (1.10) | 11 (2.39) | 51 (2.40) |
| sickle cell disease, HIV or<br>severe combined<br>immunodeficiency | 2682 (0.25) | 383 (0.26) | * | 17 (0.80) |

**SUPPLEMENTARY TABLE 2:**
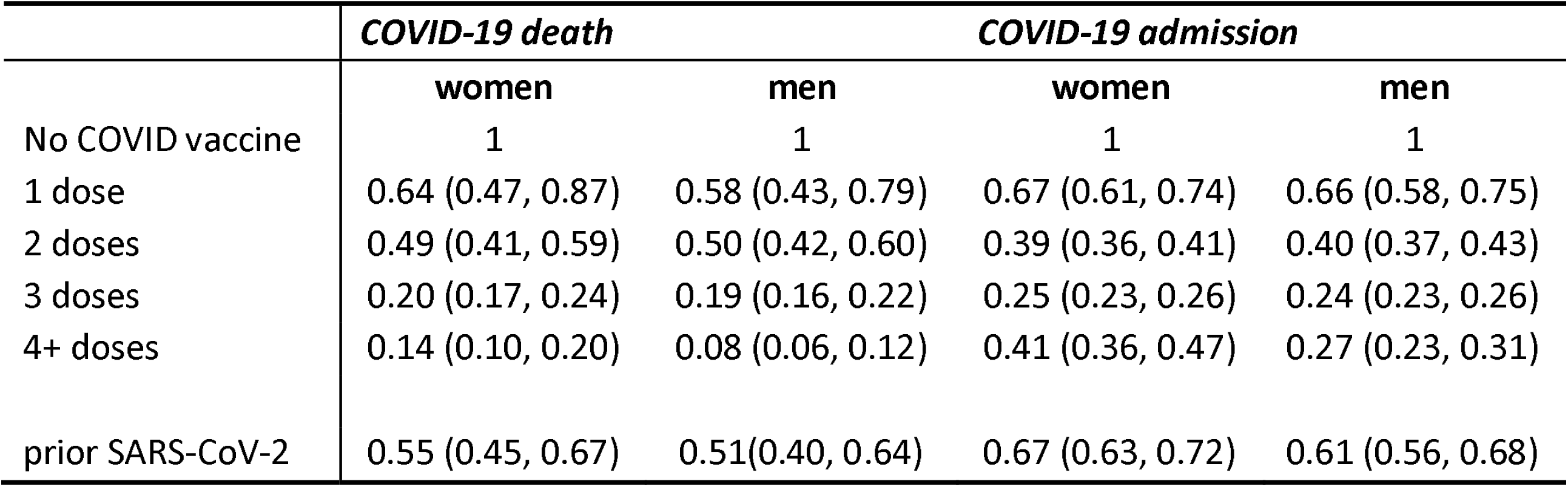
Fully adjusted HR for COVID-19 death and admission in men and women summarising the results for vaccination dose and SARS-CoV-2 infection prior to study period. HR adjusted for age and BMI as well as variables shown in the separate Figures 1-4.

**SUPPLEMENTARY TABLE 3:** Performance of the QCOVID4 algorithms in the validation cohort in subgroups for age group and ethnicity.

| statistic | COVID-19 death |  | COVID-19 admission |  |
| --- | --- | --- | --- | --- |
|  | females | males | females | males |
| **<70 years** |  |  |  |  |
| D statistic | 2.96 (2.07 to 3.84) | 2.56 (1.93 to 3.19) | 2.91 (2.04 to 3.78) | 2.42 (1.81 to 3.02) |
| Harrell's C | .915 (.856 to .973) | .894 (.846 to .942) | .945 (.918 to .972) | .904 (.863 to .945) |
| R2 | 67.6 (54.5 to 80.7) | 61.1 (49.3 to 72.8) | 66.9 (53.7 to 80.2) | 58.3 (46.1 to 70.4) |
| **70-79 years** |  |  |  |  |
| D statistic | 2.63 (2.11 to 3.15) | 2.06 (1.61 to 2.51) | 1.94 (1.47 to 2.41) | 1.73 (1.3 to 2.17) |
| Harrell's C | .88 (.831 to .929) | .833 (.777 to .889) | .817 (.765 to .869) | .79 (.733 to .848) |
| R2 | 62.3 (53 to 71.5) | 50.3 (39.4 to 61.1) | 47.3 (35.1 to 59.4) | 41.7 (29.5 to 54) |
| **80+ years** |  |  |  |  |
| D statistic | 1.12 (.79 to 1.46) | 1.17 (.905 to 1.44) | .941 (0.68 to 1.2) | 1.04 (.779 to 1.29) |
| Harrell's C | .673 (.63 to .717) | .713 (.675 to .752) | .664 (.621 to .707) | .698 (.659 to .736) |
| R2 | 23.2 (12.6 to 33.8) | 24.8 (16.2 to 33.3) | 17.4 (9.46 to 25.4) | 20.4 (12.4 to 28.5) |
| **White** |  |  |  |  |
| D statistic | 3.68 (3.44 to 3.91) | 3.64 (3.4 to 3.87) | 3.36 (3.12 to 3.59) | 3.44 (3.22 to 3.66) |
| Harrell's C | .963 (.948 to .979) | .969 (.96 to .978) | .964 (.955 to .973) | .968 (.961 to .976) |
| R2 | 76.4 (74.1 to 78.7) | 76 (73.6 to 78.3) | 72.9 (70.2 to 75.7) | 73.9 (71.4 to 76.4) |
| **Indian** |  |  |  |  |
| D statistic | * | 4.38 (1.16 to 7.61) | * | 3.61 (.811 to 6.41) |
| Harrell's C | .975 (.948 to 1) | .994 (.99 to .998) | .971 (.951 to .991) | .989 (.98 to .998) |
| R2 | * | 81.1 (57 to 105) | * | 74.6 (43.7 to 106) |
| **Pakistani** |  |  |  |  |
| D statistic | 3.5 (1.56 to 5.44) | * | 2.76 (.939 to 4.57) | * |
| Harrell's C | .971 (.938 to 1) | .989 (.983 to .995) | .867 (.668 to 1.07) | .981 (.974 to .989) |
| R2 | 74.5 (53.5 to 95.5) | * | 64.4 (34 to 94.7) | * |
| **Bangladeshi** |  |  |  |  |
| D statistic | 4.44 (.539 to 8.35) | 4.79 (.392 to 9.19) | 4.31 (.342 to 8.27) | 144 (-91.7 to 380) |
| Harrell's C | .992 (.987 to .998) | .994 (.989 to .999) | .984 (.977 to .992) | * |
| R2 | 82.4 (57.4 to 107) | 84.6 (60.4 to 109) | 81.2 (53.4 to 109) | 97.6 (81.4 to 114) |
| **Other Asian** |  |  |  |  |
| D statistic | * | 3.88 (1.37 to 6.39) | * | 5.57 (1.82 to 9.32) |
| Harrell's C | * | .974 (.951 to .998) | * | .981 (.956 to 1.01) |
| R2 | * | 78.1 (55.1 to 101) | * | 87.8 (74 to 102) |
| **Caribbean** |  |  |  |  |
| D statistic | 5.6 (.00527 to 11.2) | 4.64 (2.12 to 7.17) | * | 3.81 (1.91 to 5.71) |
| Harrell's C | .997 (.993 to 1) | .969 (.947 to .991) | .984 (.974 to .993) | .965 (.944 to .986) |
| R2 | 87.9 (68.1 to 108) | 83.6 (68.9 to 98.3) | 76.3 (41.1 to 112) | 77.5 (60.4 to 94.7) |
| **Black african** |  |  |  |  |
| D statistic | 4.93 (1.77 to 8.1) | 41.6 (-196 to 279) | 4.34 (1.5 to 7.18) | 5.69 (1.06 to 10.3) |
| Harrell's C | .995 (.992 to .998) | .995 (.989 to 1) | .993 (.988 to .997) | .998 (.995 to 1) |
| R2 | 85.3 (69.2 to 101) | 92.9 (76.6 to 109) | 81.8 (62.4 to 101) | 88.4 (71.6 to 105) |
| **Chinese** |  |  |  |  |
| D statistic | 3.06 (-5.39 to 11.5) | 4.18 (-.867 to 9.22) | 2.93 (-5.05 to 10.9) | 4.17 (-1.19 to 9.52) |
| Harrell's C | * | .993 (.985 to 1) | * | .988 (.978 to .999) |
| R2 | 51.4 (-90.9 to 194) | 76.9 (23.7 to 130) | * | 76 (17.5 to 135) |
| **Other** |  |  |  |  |
| D statistic | 5.74 (2.15 to 9.33) | 3.17 (1.57 to 4.78) | 5.25 (1.8 to 8.7) | 3.2 (1.53 to 4.87) |
| Harrell's C | .994 (.985 to 1) | .945 (.903 to .986) | .992 (.98 to 1) | .929 (.866 to .992) |
| R2 | 88.6 (76.3 to 101) | 70.5 (49.2 to 91.8) | 86.5 (71.9 to 101) | 70.7 (48.7 to 92.7) |

\*counts too low for analysis

**SUPPLEMENTARY TABLE 4:** characteristics of 3,600 patients in the NHS Digital high-risk cohort and 36000 patients with the highest predicted risks of COVID-19 mortality using QCOVID4 in the validation cohort.

|  | *validation cohort* | *High Risk*<br>*QCOVID4* | *High risk NHS*<br>*digital* |
| --- | --- | --- | --- |
| Total number | 145397 | 3600 | 3600 |
| Men | 62305 (42.85) | 1682 (46.72) | 1500 (41.67) |
| COVID-19 deaths | 461 (0.32) | 333 (9.25) | 95 (2.64) |
| COVID-19 admissions | 2873 (1.98) | 815 (22.64) | 462 (12.83) |
| Age (SD) | 42.96 (16.41) | 85.00 (7.30) | 55.40 (17.87) |
| Town (SD) | -0.01 (3.03) | -0.15 (2.94) | -0.05 (3.04) |
| Body mass index (SD) | 26.62 (5.67) | 25.69 (5.87) | 27.86 (6.06) |
| 20-29 years | 34453 (23.70) | - | 286 (7.94) |
| 30-39 years | 35586 (24.48) | - | 495 (13.75) |
| 40-49 years | 28922 (19.89) | - | 596 (16.56) |
| 50-59 years | 22507 (15.48) | 11 (0.31) | 692 (19.22) |
| 60-69 years | 12745 (8.77) | 97 (2.69) | 663 (18.42) |
| 70-79 years | 7056 (4.85) | 653 (18.14) | 523 (14.53) |
| 80-89 years | 4128 (2.84) | 2839 (78.86) | 345 (9.58) |
| White | 123145 (84.70) | 3341 (92.81) | 3006 (83.50) |
| Indian | 3523 (2.42) | 54 (1.50) | 59 (1.64) |
| Pakistani | 2831 (1.95) | 48 (1.33) | 78 (2.17) |
| Bangladeshi | 1894 (1.30) | 26 (0.72) | 47 (1.31) |
| Other Asian | 2476 (1.70) | 19 (0.53) | 36 (1.00) |
| Caribbean | 1142 (0.79) | 40 (1.11) | 52 (1.44) |
| Black african | 3256 (2.24) | 35 (0.97) | 160 (4.44) |
| Chinese | 1128 (0.78) | 11 (0.31) | 32 (0.89) |
| Other ethnic group | 6002 (4.13) | 26 (0.72) | 130 (3.61) |
| no CKD | 141747 (97.49) | 2244 (62.33) | 3017 (83.81) |
| CKD3 | 3153 (2.17) | 1179 (32.75) | 282 (7.83) |
| CKD4 | 167 (0.11) | 96 (2.67) | 37 (1.03) |
| CKD5 only | 164 (0.11) | 59 (1.64) | 110 (3.06) |
| CKD5 with dialysis | 53 (0.04) | 5 (0.14) | 41 (1.14) |
| CKD5 with transplant | 113 (0.08) | 17 (0.47) | 113 (3.14) |
| no learning disability | 142758 (98.18) | 3496 (97.11) | 3443 (95.64) |
| Learning disability | 2553 (1.76) | 102 (2.83) | 72 (2.00) |
| Downs syndrome | 86 (0.06) | * | 85 (2.36) |
| No chemo in last 12 | 144888 (99.65) | 3435 (95.42) | 3342 (92.83) |
| Chemo group A | 207 (0.14) | 53 (1.47) | 31 (0.86) |
| chemo group B | 284 (0.20) | 106 (2.94) | 212 (5.89) |
| chemo group C | 18 (0.01) | 6 (0.17) | 15 (0.42) |
| no covid vaccine doses | 15725 (10.82) | 211 (5.86) | 161 (4.47) |
| 1 dose | 4985 (3.43) | 82 (2.28) | 67 (1.86) |
| 2 doses | 39637 (27.26) | 513 (14.25) | 485 (13.47) |
| 3 doses | 84272 (57.96) | 2735 (75.97) | 2567 (71.31) |
| 4 doses | 778 (0.54) | 59 (1.64) | 320 (8.89) |
| no therapeutics | 144564 (99.43) | 3469 (96.36) | 3096 (86.00) |
| Tocilizumab | 54 (0.04) | 13 (0.36) | 14 (0.39) |
| Sotrovimab | 406 (0.28) | 50 (1.39) | 292 (8.11) |
| sarilumab | 22 (0.02) | 7 (0.19) | * |
| Casirivimab and Imdevimab | 20 (0.01) | 10 (0.28) | 8 (0.22) |
| Remdesivir | 108 (0.07) | 29 (0.81) | 24 (0.67) |
| Molnupiravir | 110 (0.08) | 13 (0.36) | 95 (2.64) |
| Paxlovid | 113 (0.08) | 9 (0.25) | 68 (1.89) |
| SARS-CoV-2 infection prior to study | 16014 (11.01) | 199 (5.53) | 333 (9.25) |
| Blood cancer | 998 (0.69) | 216 (6.00) | 526 (14.61) |
| Bone marrow transplant in<br>previous 6 months | * | * | * |
| Respiratory cancer | 211 (0.15) | 74 (2.06) | 38 (1.06) |
| radiotherapy in last 6 months | 181 (0.12) | 54 (1.50) | 106 (2.94) |
| solid organ transplant ever | 49 (0.03) | 7 (0.19) | 46 (1.28) |
| COPD | 1889 (1.30) | 580 (16.11) | 331 (9.19) |
| pulmonary hypertension | 87 (0.06) | 31 (0.86) | 23 (0.64) |
| coronary heart disease | 3312 (2.28) | 935 (25.97) | 290 (8.06) |
| stroke | 2020 (1.39) | 675 (18.75) | 175 (4.86) |
| atrial fibrillation | 2457 (1.69) | 955 (26.53) | 234 (6.50) |
| congestive cardiac failure | 1251 (0.86) | 581 (16.14) | 159 (4.42) |
| VTE | 2269 (1.56) | 392 (10.89) | 236 (6.56) |
| peripheral vascular disease | 586 (0.40) | 235 (6.53) | 56 (1.56) |
| congenital heart disease | 821 (0.56) | 13 (0.36) | 45 (1.25) |
| dementia | 1630 (1.12) | 1100 (30.56) | 103 (2.86) |
| parkinson's disease | 246 (0.17) | 129 (3.58) | 16 (0.44) |
| epilepsy | 1911 (1.31) | 106 (2.94) | 103 (2.86) |
| rare neurological conditions | 465 (0.32) | 27 (0.75) | 386 (10.72) |
| cerebral palsy | 163 (0.11) | 2 (0.06) | 8 (0.22) |
| osteoporotic fracture | 5429 (3.73) | 731 (20.31) | 282 (7.83) |
| RA or SLE | 3310 (2.28) | 348 (9.67) | 365 (10.14) |
| cirrhosis | 241 (0.17) | 40 (1.11) | 139 (3.86) |
| bipolar disorder or schizophrenia | 1392 (0.96) | 58 (1.61) | 42 (1.17) |
| inflammatory bowel disease | 1600 (1.10) | 57 (1.58) | 280 (7.78) |
| sickle cell disease, HIV or severe<br>combined immunodeficiency | 383 (0.26) | 12 (0.33) | 349 (9.69) |
| type 1 diabetes | 844 (0.58) | 16 (0.44) | 56 (1.56) |
| type 2 diabetes | 6322 (4.35) | 997 (27.69) | 473 (13.14) |

## LEGENDS FOR FIGURES

**Supplementary Figure 1a Adjusted hazard ratio (95% CI) by age for risk of COVID-19 mortality**

**Supplementary Figure 1b Adjusted hazard ratio (95% CI) by BMI for COVID-19 mortality**

**Supplementary Figure 1c Adjusted hazard ratio (95% CI) by age for risk of COVID-19 admission**

**Supplementary Figure 1d Adjusted hazard ratio (95% CI) by BMI for COVID-19 admission**

**Supplementary Figure 2 Calibration of the QCOVID2 risk model to predict COVID-19 death following vaccination**

**Supplementary Figure 3 Calibration of the QCOVID2 risk model to predict COVID-19 admission following vaccination**

## Notes

### Competing Interest Statement

All authors have completed the ICMJE uniform disclosure form. JHC reports grants from National Institute for Health Research (NIHR) Biomedical Research Centre, Oxford, grants from John Fell Oxford University Press Research Fund, grants from Cancer Research UK (CR-UK) grant number C5255 A18085, through the Cancer Research UK Oxford Centre, grants from the Oxford Wellcome Institutional Strategic Support Fund and other research councils, during the conduct of the study. JHC is an unpaid director of QResearch, a not-for-profit organisation which is a partnership between the University of Oxford and EMIS Health who supply the QResearch database used for this work. JHC is a founder and shareholder of ClinRisk ltd and was its medical director until 31st May 2019. ClinRisk Ltd produces open and closed source software to implement clinical risk algorithms (outside this work) into clinical computer systems. JHC is chair of the NERVTAG risk stratification subgroup and a member of SAGE COVID-19 groups and the NHS group advising on prioritisation of use of monoclonal antibodies in COVID-19 infection. CC reports receiving personal fees from ClinRisk Ltd, outside this work and is a member of the NERVTAG risk stratification subgroup. KK is Chair of the Ethnic Subgroup of the UK Scientific Advisory Group for Emergencies (SAGE) and a member of SAGE. AS has received research grants from NIHR, MRC, HDRUK, CSO, NCS, GSK for COVID-19 work and has provided advice for AstraZeneca Thrombotic Thrombocytopenic Taskforce and the UK and Scottish Government COVID-19 Advisory Groups (roles unremunerated)

### Author Declarations

The QResearch ethics approval was provided on 8th June 2020 by the East Midlands-Derby Research Ethics Committee reference 18 EM 0400.

## REFERENCES

1. Clift AK, Coupland CAC, Keogh RH, et al. Living risk prediction algorithm (QCOVID) for risk of hospital admission and mortality from coronavirus 19 in adults: national derivation and validation cohort study. BMJ 2020;371:m3731. doi: 10.1136/bmj.m3731 [published Online First: 2020/10/22]

2. Nafilyan V, Humberstone B, Mehta N, et al. An external validation of the QCovid risk prediction algorithm for risk of mortality from COVID-19 in adults: a national validation cohort study in England. The Lancet Digital Health 2021 doi: 10.1016/s2589-7500(21)00080-7

3. Lyons J NV, Akbari A, Davies G, Griffiths Rowena, Harrison E, Hippisley-Cox J, Hollinghurst J, Khunti K, North L, Sheikh A, Torabi F, Lyons R.. Validating the QCOVID risk prediction algorithm for risk of mortality from COVID-19 in the adult population in Wales, UK. International Journal of Population Data Science 2022

4. Simpson CR, Robertson C, Kerr S, et al. External validation of the QCovid risk prediction algorithm for risk of COVID-19 hospitalisation and mortality in adults: national validation cohort study in Scotland. Thorax 2021 doi: 10.1136/thoraxjnl-2021-217580

5. NHS Digital. Coronavirus Shielded Patient List open data set, England 2021 [cited 2021 01/0/07/2021]. Available from: https://digital.nhs.uk/dashboards/shielded-patient-list-open-data-set.

6. Hippisley-Cox J, Coupland CA, Mehta N, et al. Risk prediction of covid-19 related death and hospital admission in adults after covid-19 vaccination: national prospective cohort study. BMJ 2021;374:2244. doi: 10.1136/bmj.n2244 [published Online First: 20210917]

7. Ward IL, Bermingham C, Ayoubkhani D, et al. Risk of covid-19 related deaths for SARS-CoV-2 omicron (B.1.1.529) compared with delta (B.1.617.2): retrospective cohort study. BMJ 2022;378:e070695. doi: 10.1136/bmj-2022-070695

8. Gupta A, Gonzalez-Rojas Y, Juarez E, et al. Early Treatment for Covid-19 with SARS-CoV-2 Neutralizing Antibody Sotrovimab. N Engl J Med 2021;385(21):1941–50. doi: 10.1056/NEJMoa2107934 [published Online First: 20211027]

9. Department of Health and Social Care. Interim Clinical Commissioning Policy: Neutralising monoclonal antibodies or antivirals for non-hospitalised patients with COVID-19 (C1530) 2021 [cited 2022 09/01/2022]. Available from: https://www.england.nhs.uk/coronavirus/publication/interim-clinical-commissioning-policy-neutralising-monoclonal-antibodies-or-antivirals-for-non-hospitalised-patients-with-covid-19/ accessed 09/01/2022 2022.

10. NHS Digital. Coronavirus Treatments 2021 [Available from: https://digital.nhs.uk/coronavirus/treatments#top.

11. McInnes I GC, Beale R, Hippisley-Cox J Barnes E, Lowe D, Misbah S, Schmid M, Screaton G< Semple C, Underwood M, Wedderburn L, Whittaker E, Snape M, de Silver T, Moss etc al. Higher-risk patients eligible for COVID-19 treatments: independent advisory group report: Department of Health and Social care, 2022.

12. Clift AK CCKRHH, Hippisley-Cox J. COVID-19 Mortality Risk in Down Syndrome: Results From a Cohort Study Of 8 Million Adults. Annals of Internal Medicine 2020;0(0):null. doi: 10.7326/m20-4986 %m 33085509

13. Hippisley-Cox J, Young D, Coupland C, et al. Risk of severe COVID-19 disease with ACE inhibitors and angiotensin receptor blockers: cohort study including 8.3 million people. Heart 2020 doi: 10.1136/heartjnl-2020-317393 [published Online First: 2020/08/02]

14. Hippisley-Cox J, Coupland C. Risk of myocardial infarction in patients taking cyclo-oxygenase-2 inhibitors or conventional non-steroidal anti-inflammatory drugs: Population based nested case-control analysis. BMJ 2005;330(7504):1366–69. doi: 10.1136/bmj.38456.398507.8F

15. UK Government. COVID-19 deaths in the UK: Uk Government; 2021 [accessed 10 Aug 2021 2021.

16. Hippisley-Cox J, Clift AK, Coupland CAC, et al. Protocol for the development and evaluation of a tool for predicting risk of short-term adverse outcomes due to COVID-19 in the general UK population. medRxiv 2020:2020.06.28.20141986-2020.06.28.86. doi: 10.1101/2020.06.28.20141986

17. Gao M, Piernas C, Astbury NM, et al. Associations between body-mass index and COVID-19 severity in 6·9 million people in England: a prospective, community-based, cohort study. The Lancet Diabetes & Endocrinology 2021 doi: 10.1016/s2213-8587(21)00089-9

18. Townsend P, Davidson N. The Black report. London: Penguin 1982.

19. Little RJA, Rubin DB. Statistical analysis with missing data/Roderick J.A. Little, Donald B. Rubin. Hoboken, N.J: Wiley, c2002. 2nd ed 2002.

20. Hosmer D, Lemeshow S. Applied Logistic Regressopm. New York: John Wiley & Sons, Inc. 1989.

21. Harrell F, Lee K, Mark D. Multivariable prognostic models: issues in developing models, evaluating assumptions and adequacy, and measuring and reducing errors. Stat Med 1996;15:361 – 87.

22. Royston P. Explained variation for survival models. Stata J 2006;6:1–14.

23. Coviello V, Boggess M. Cumulative incidence estimation in the presence of competing risks. The Stata Journal 2004;4(2):103–12.

24. Benchimol EI, Smeeth L, Guttmann A, et al. The REporting of studies Conducted using Observational Routinely-collected health Data (RECORD) statement. PLoS Med 2015;12(10):e1001885. doi: 10.1371/journal.pmed.1001885 [published Online First: 2015/10/07]

25. Collins GS, Reitsma JB, Altman DG, et al. Transparent Reporting of a multivariable prediction model for Individual Prognosis Or Diagnosis (TRIPOD): The TRIPOD StatementThe TRIPOD Statement. Ann Int Med 2015;162(1):55–63. doi: 10.7326/M14-0697

26. Care CSfHaS. Citizens’ Jury on QCovid: Report on the jury’s conclusions and key findings: Scottish Government; 2022 [Available from: https://www.gov.scot/publications/citizens-jury-qcovid-report-jurys-conclusions-key-findings/pages/3/ accessed 03/08/2022 2022.

27. <bmj-2022-070695.full.pdf>. doi: 10.1136/bmj-2022-070695

28. Hippisley-Cox J, Coupland C, Brindle P. Development and validation of QRISK3 risk prediction algorithms to estimate future risk of cardiovascular disease: prospective cohort study. BMJ 2017;357:j2099. doi: 10.1136/bmj.j2099

29. Hippisley-Cox J, Coupland C. Development and validation of QDiabetes-2018 risk prediction algorithm to estimate future risk of type 2 diabetes: cohort study. BMJ 2017;359:j5019. doi: 10.1136/bmj.j5019

30. Hippisley-Cox J, Coupland C. Development and validation of QMortality risk prediction algorithm to estimate short term risk of death and assess frailty: cohort study. BMJ 2017;358:j4208. doi: 10.1136/bmj.j4208 [published Online First: 2017/09/22]

31. Collins GS, Altman DG. An independent external validation and evaluation of QRISK cardiovascular risk prediction: a prospective open cohort study. BMJ 2009;339:b2584.. doi: 10.1136/bmj.b2584

32. Collins GS, Altman DG. An independent and external validation of QRISK2 cardiovascular disease risk score: a prospective open cohort study. BMJ 2010;340:c2442. doi: 10.1136/bmj.c2442

33. Collins GS, Altman DG. Predicting the 10 year risk of cardiovascular disease in the United Kingdom: independent and external validation of an updated version of QRISK2. BMJ 2012;344:e4181. doi: 10.1136/bmj.e4181

34. Nafilyan V, Islam N, Mathur R, et al. Ethnic differences in COVID-19 mortality during the first two waves of the Coronavirus Pandemic: a nationwide cohort study of 29 million adults in England. Eur J Epidemiol 2021;36(6):605–17. doi: 10.1007/s10654-021-00765-1 [published Online First: 20210616]

