## Supplementary figures and images for "QCovid 4 - Predicting risk of death or hospitalisation from COVID-19 in adults testing positive for SARS-CoV-2 infection during the Omicron wave in England"

### Supplementary figure 1

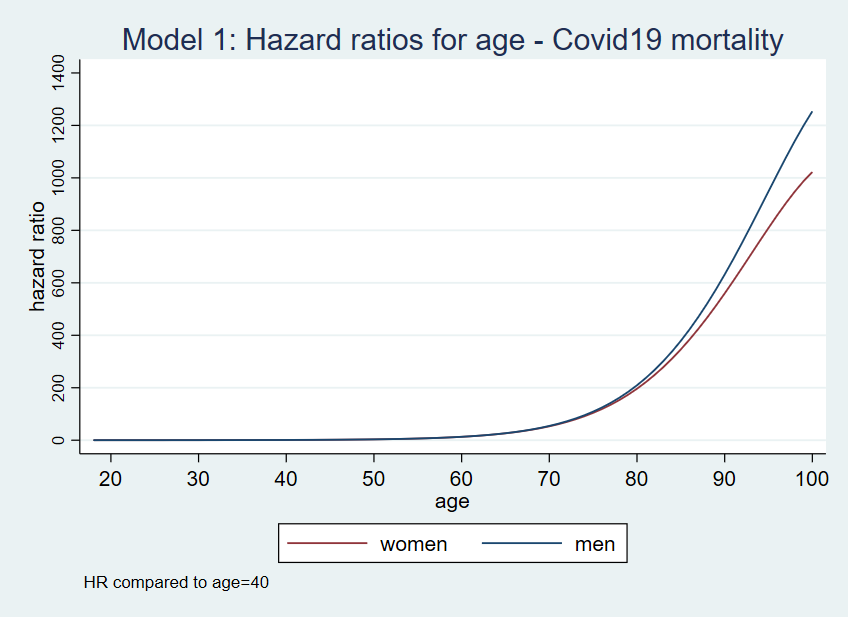

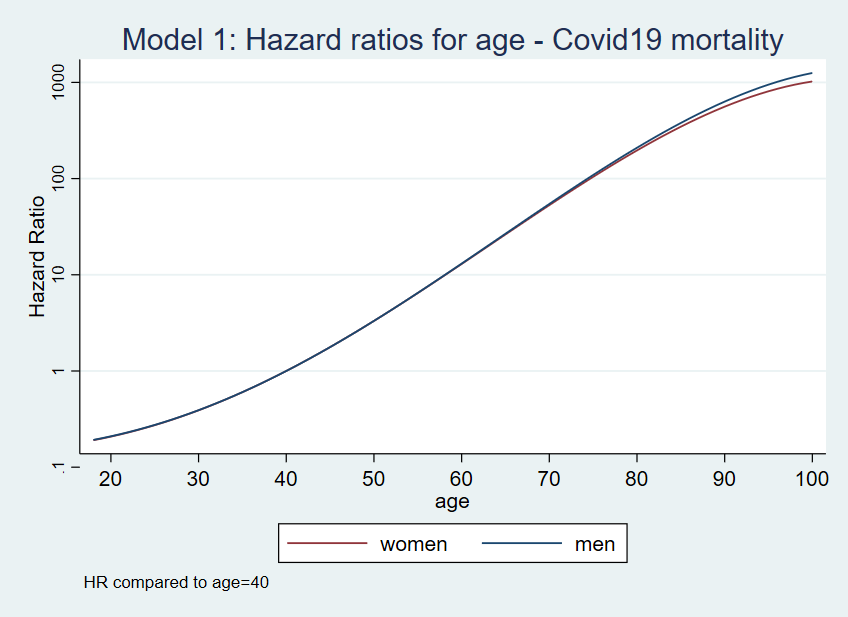

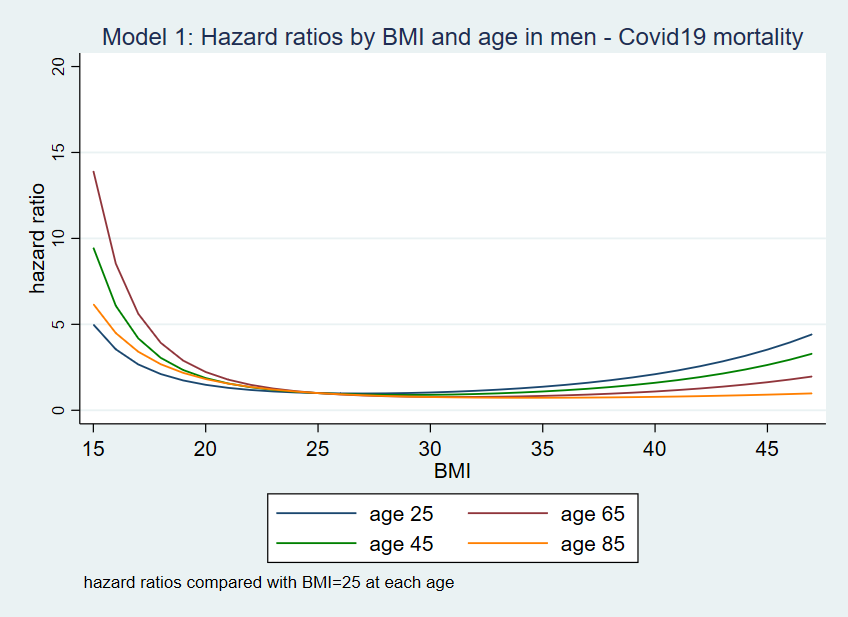

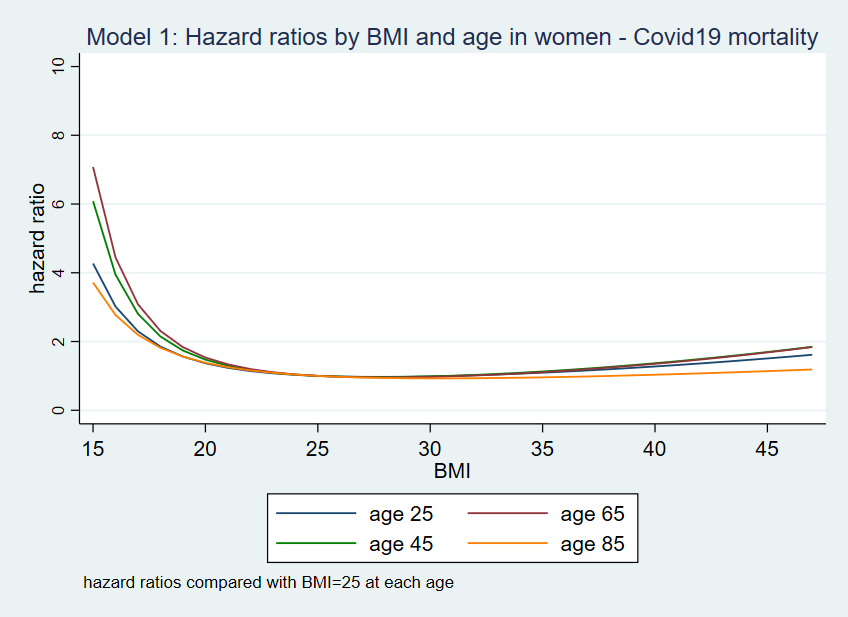

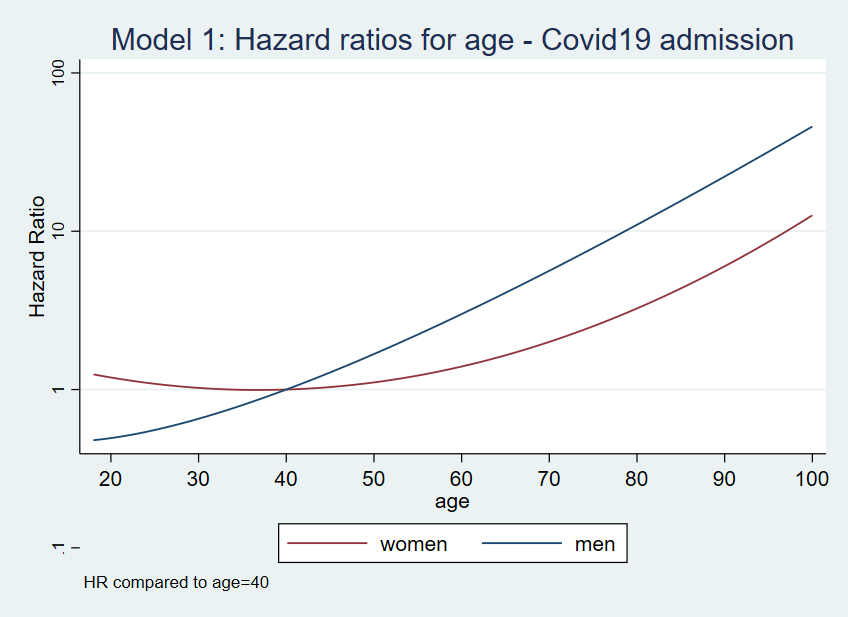

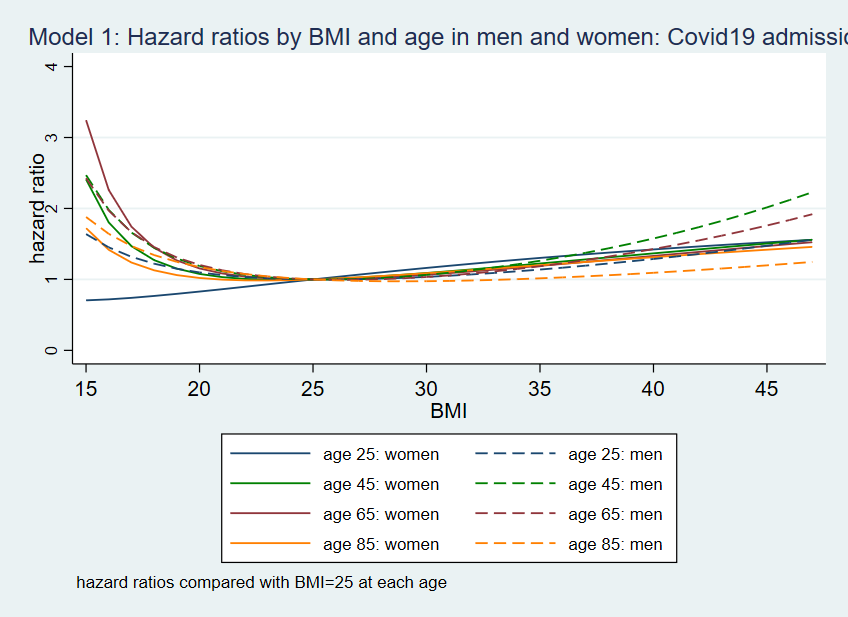

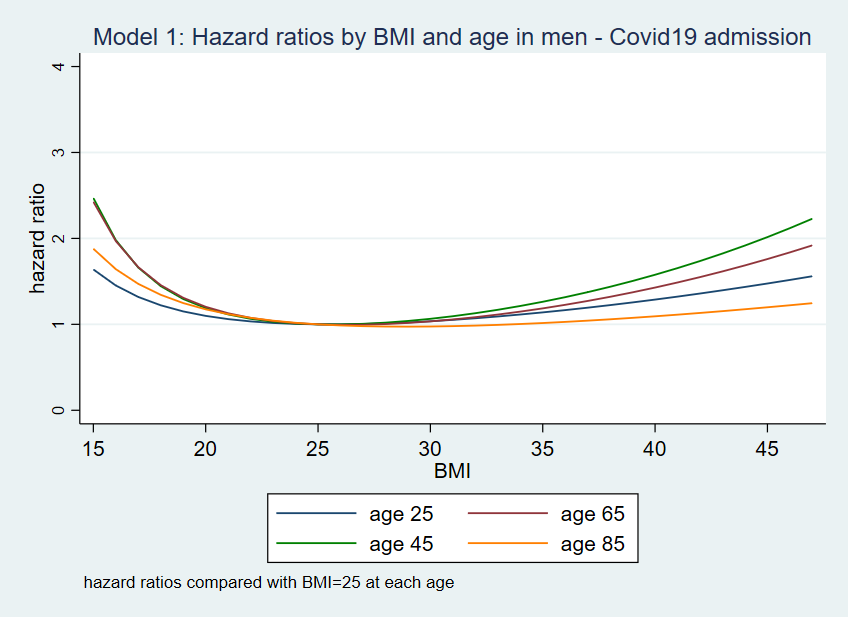

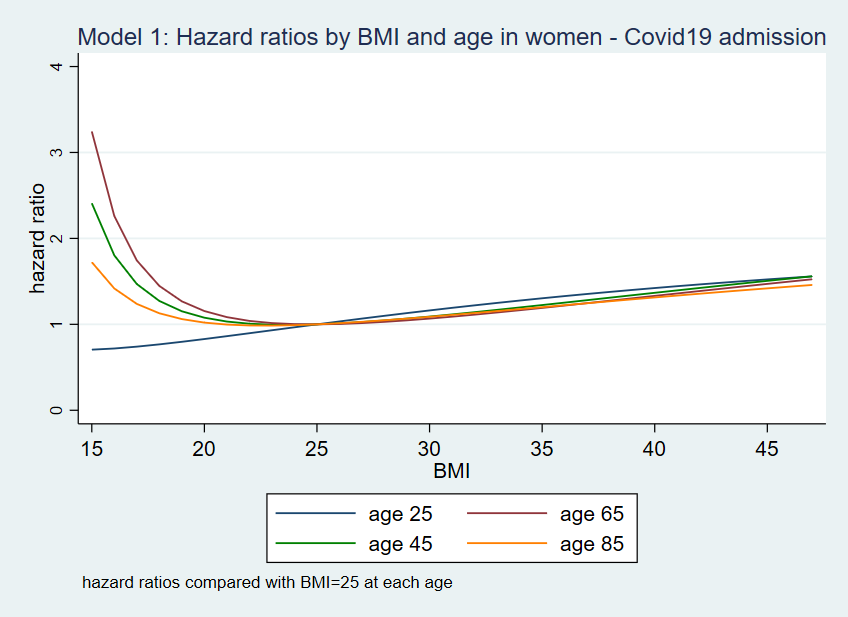

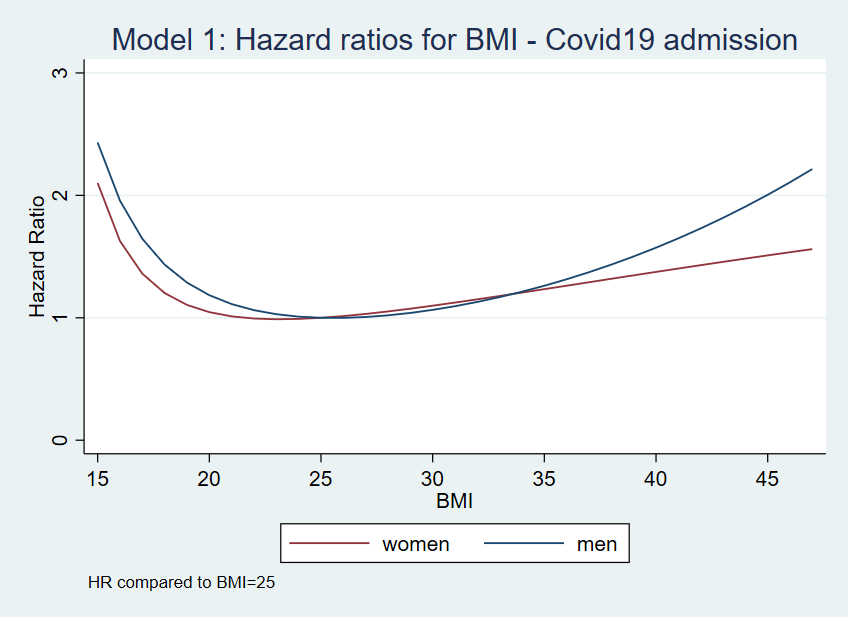

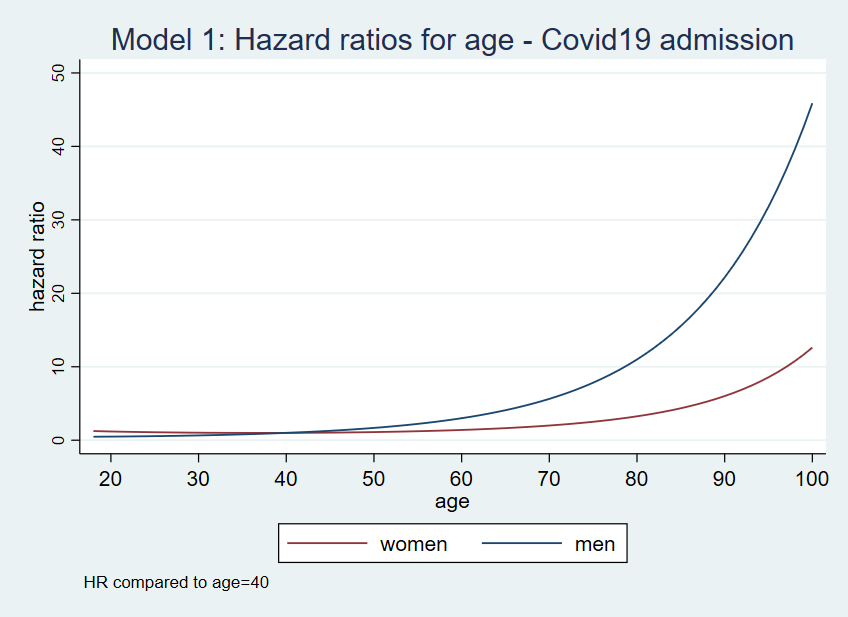

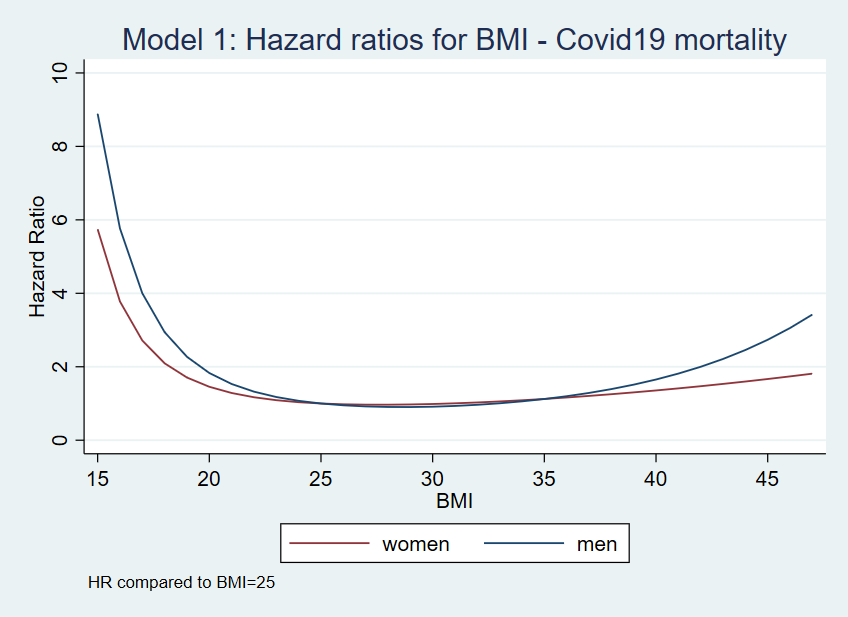
